# Assessing the Intervention’s Effectiveness and Health System Efficiency During COVID-19 Crisis using A Signal-to-Noise Ratio Index

**DOI:** 10.1101/2020.05.07.20094334

**Authors:** Khaled Wahba

## Abstract

During COVID-19 nearly everyone around the globe was monitoring the situation on a daily, if not hourly, basis by tracking a set of numbers that were reported by different institutions through multiple platforms: either official, or informal. Irrespective of the sources from which the data was pulled, many researchers, reporters and professionals made the effort to represent the data in different ways in an effort to explain: what happened, what was happening, and what might happen; with the hope of seeing a sign of slowing down the spread of the virus (SARS-CoV-2). A subset of these reported numbers included: the confirmed cases, number of deaths, number of recovered cases along with the number of tests being carried out by each country. Each of these numbers (metrics) was able to reveal only one side of the reality ignoring the messages that might come from other metrics (numbers). Focusing only on one single metric to reflect on the situation opened the doors for emergent opinions, theories speculations, and even confusion among the professionals before the public. In fact, all of these efforts to explain and describe the situation through the same available numbers did not manage to see clearly or shed the light on the performance and the efficiency of the country’s health system in dealing with the ongoing COVID-19 outbreak. It was evident that none of these numbers could reconstruct the full picture about the virus spread behavior nor about the capability and capacity of the health system in dealing with the pandemic. A combined metric should have been developed to best reflect the performance of the health system during the crisis. In this paper, a signal to noise ratio like index, *snr* was introduced in an attempt to evaluate the efficiency of the health system as well as the effectiveness of the interventions taken by the stakeholders in an effort to control the virus spread during any health-related crisis. Using this proposed index (*snr*), it was possible to carry out a data-driven comparison among different countries in their efforts of dealing with the crisis. The primary focus of this study was to assess the interventions’ effectiveness by the decision makers along with the health system’s efficiency of the countries that experienced a relatively high pressure and stress on their systems. In this study, 19 countries were selected based on predefined criteria that included: (1) the reported total confirmed cases should exceed 5,000, and (2) the total confirmed cases per 1 million people should exceed 200, at the time this study was concluded.

According to the proposed *snr* index, the findings showed that Germany and South Korea were ahead of the game, by far, compared to other countries such as the USA, Spain, Italy, Belgium, Netherlands, and UK. Some other countries, such as Canada, Austria, Switzerland managed to slightly pivot their interventions at a later stage in effective manners according to the *snr* index. The study explains the foundation, and the underlying calculations of the proposed *snr* index. Moreover, the study shows how reliably the *snr* index measures the interventions’ effectiveness and health system’s efficiency during the crisis or during any health related crisis.

An additional, yet interesting finding from this study, was that the *snr* curve showed a persistent four episode (segment) structure or pattern during the pandemic. This finding could suggest a benchmark of the expected pattern of the fight against the virus spread during the pandemic that could offer a significant tool, or approach, for the decision makers.

Finally, it is worth mentioning that the implementation of this proposed index is only valid and meaningful during a crisis. In a none crisis time, the required data to calculate the *snr* index is not available and rather mathematically misleading.

## 1 Introduction

Human beings, since creation, have been going through different and tough crises ranging from nature, environmental, economic, financial, political, technological and health related outbreaks. All of them have had extremely negative impacts on the way human beings survived after, which led to a series of fundamental disruptions on all facets of human life. The COVID-19 pandemic that started in China by the end of December 2019 and spread around the world causing a lot deaths is not the first, and will not be the last outbreak, that humans will encounter. Figure 1 lists the most deadly pandemics occurred along the human beings history [1].

**Figure 1:**
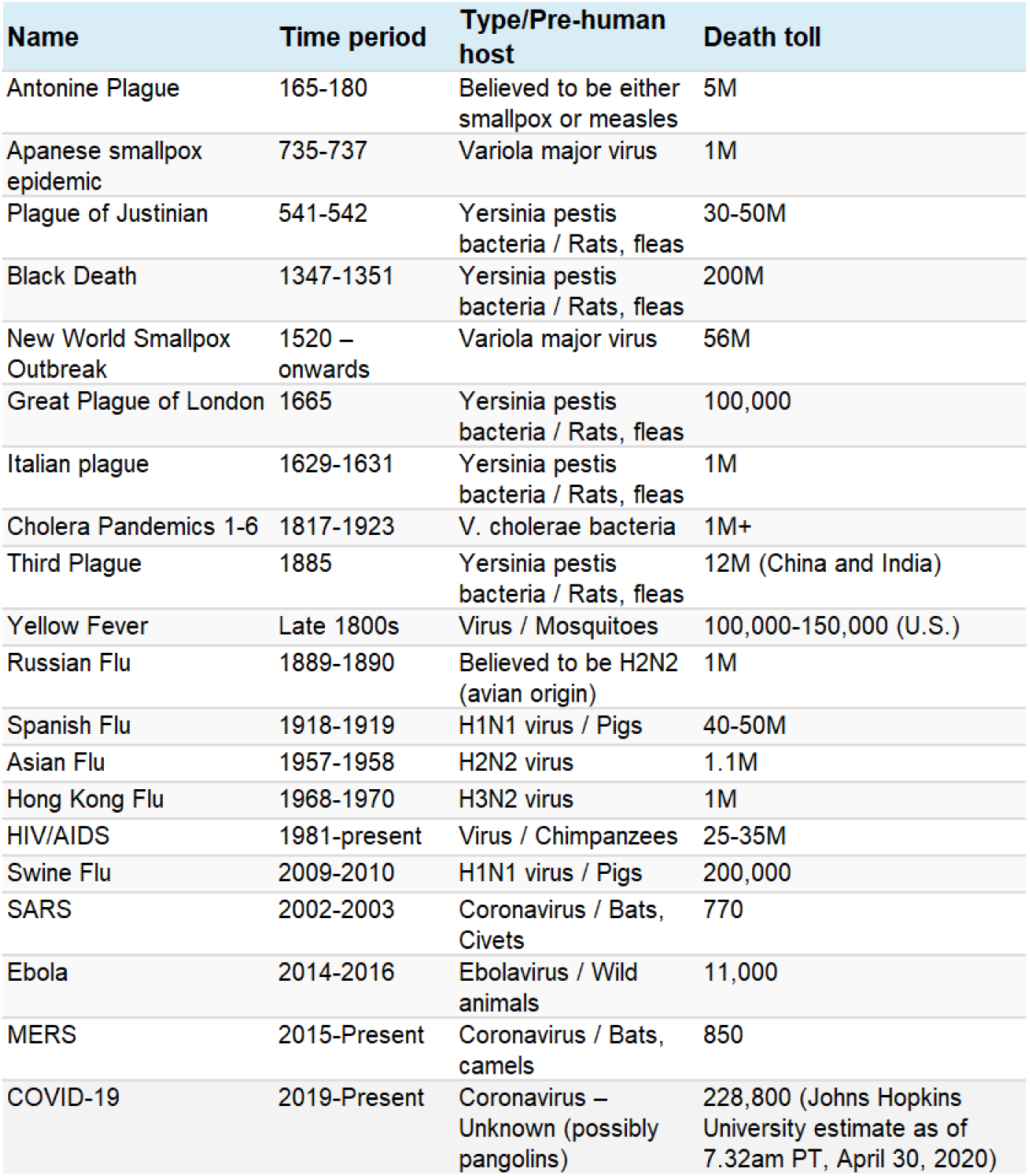
List of the most deadly pandemics occurred along the human beings history.

The extent of the losses caused by a pandemic has been dependent on several critical factors that include but are not limited to: the severity of the virus, the resilience degree of the immune system of the infected people, and the readiness of the operating systems in facing the pandemic. All operating systems must be at a high degree of integration, alignments and cooperation during the crisis. They include: healthcare systems, political systems, economic and financial systems, legal systems, information and telecommunication systems, social system, environmental systems, and technological systems. Surviving a pandemic with the least losses requires different types of management. For this kind of crisis, swift actions and interventions by the decision makers are of the utmost priority in facing the unpredictable behavior of the outbreak.

Discussing the related medical issues of the COVID-19 outbreak is out of scope for this study. The main purpose of this paper is to discuss the managerial aspects that are associated with the pandemic management. A successful crisis management practice requires the team to accurately identify the course of actions (interventions) and properly define the key performance indicators (KPI) to measure the effect of the interventions. Due to the chaotic nature of the pandemic, the team should be proactively prepared with a set of agreed upon predefined actions that are related to health, economic, legal, social, educational, environmental, and political aspects.

During any pandemic, various indicators are used and continuously reported to monitor the development of the situation. These indicators include, but are not limited to [2]: the case fatality rate (CFR), the crude mortality rate (or crude death rate), number of tests carried out, and number of infected people. The case fatality rate (CFR) is calculated through dividing total number of deaths by the total number of infected people (the diagnosed cases only). This measure does not reflect the true mortality rate, or the infection fatality rate (IFR), due to a lack of accurate data collection regarding the true number of positive cases that are diagnosed and not diagnosed. Another not unimportant indicator is the crude mortality rate or crude death rate which is calculated through dividing the number of deaths by the total population.

### 1.1 Interventions and government responses during the pandemic

To slow down the spread of a pandemic, governments all over the world had to take drastic measures that addressed health, economic and social aspects within their country. A list of these interventions and responses was reported by different institutions such as university of Oxford that included [3]: school closing, workplace closing, cancel public events, close public transport, public information campaigns, restrictions on internal movement, and international travel controls. On other side, ACAPS [4], classified the interventions into five categories: social distancing, movement restrictions, public health measures, social and economic measures, and lockdowns. In this study, ACAPS’s classification was used due to the completeness of its dataset regarding other metrics that are timely synchronized and required for further analysis. The dataset used in this study was compiled by Gassen [5, 6].

### 1.2 Healthcare system capacity: Worldwide view

Healthcare system efficiency, in terms of definitions and measurements, has been a serious and critical subject for scholars and professionals [7, 8, 9]. In simple words, the efficiency of a system could be defined as the ratio between the gained value from the system (outputs) and the amount of consumed resources (inputs) to deliver this value. For a complex system; like healthcare that has a high degree of inter-dependency among its components and subsystems, it is not a trivial task to quantify the gained value neither is assessing the consumed resources. Healthcare system resources; however, can be classified into tangible and intangible categories. While the tangible category (quantitative measures) includes: health workers (doctors, nurses, technical and managerial staff), hospital beds, and equipment, the intangible category (qualitative) includes: skills, knowledge and quality of the tangible (nonhuman) resources. Irrespective of which metrics, methods or frameworks are applied to assess the efficiency of the health systems; indubitably few metrics such as: number of hospital beds in 1000 people and number of doctors (or physicians) per 1000 people are not uncommon metrics for assessing the capacity of the healthcare systems in a country [10].

Figures 2 and 3, present the capacity of some selected countries over the duration from 1960 to 2014 and 2018, respectively. The countries in figure 2 can be classified into three groups: top, middle and bottom. The top group includes, with no surprise: Austria, Germany and Switzerland, while the bottom group includes countries like: Turkey, Brazil, South Korea, Canada, United states and United Kingdom. On the other side, figure 3 shows the number of hospital beds per 1,000 people indicating, that once again, Germany and Austria and South Korea are in the top group, leaving Iran, Canada, United States and United Kingdom in the bottom. These figures represent strong evidence in justifying and reflecting the performance of the health systems of the correspondent countries during the COVID-19 pandemic.

**Figure 2:**
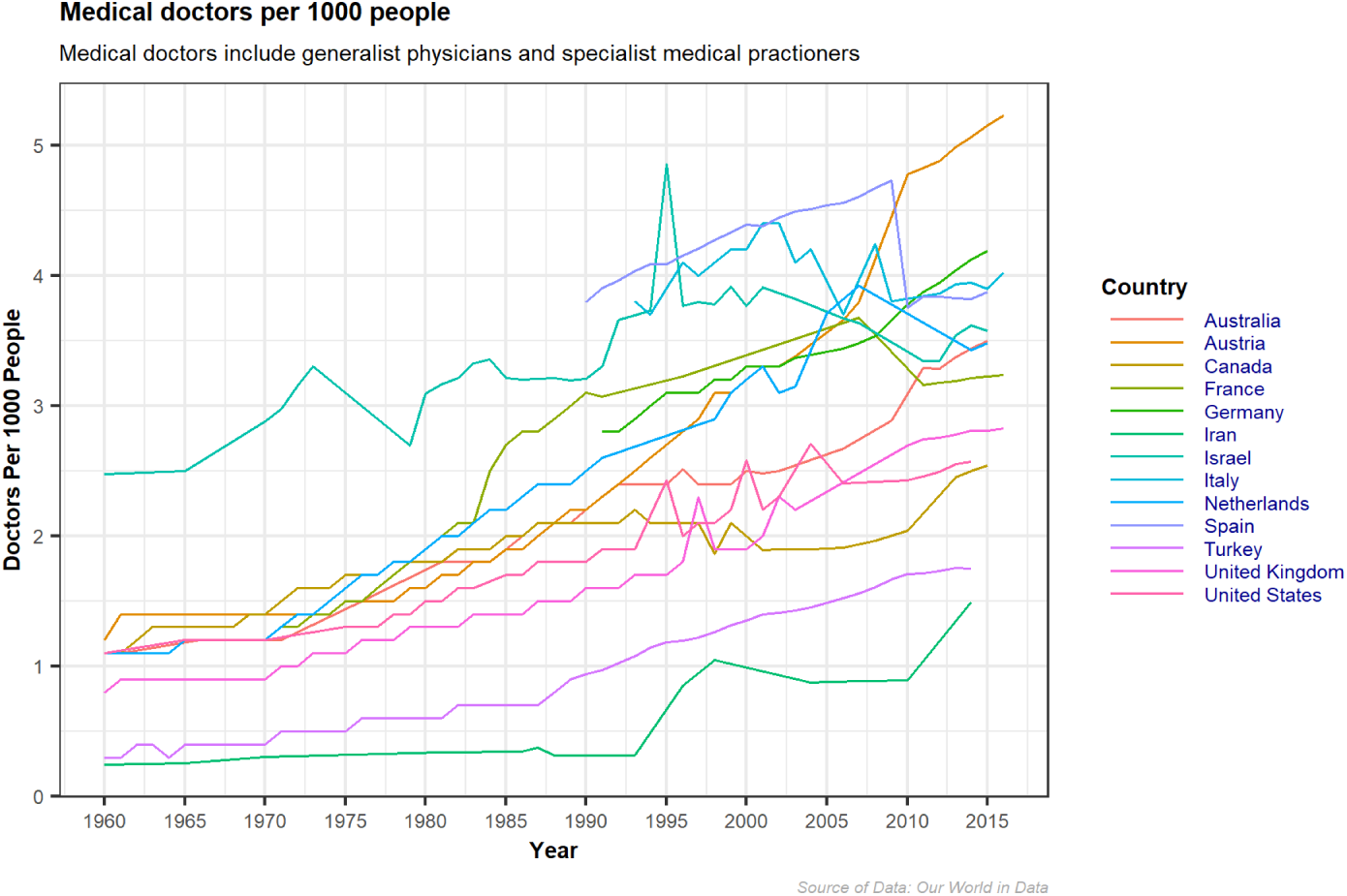
Healthcare Capacity: Medical Doctors per 1000 People.

**Figure 3:**
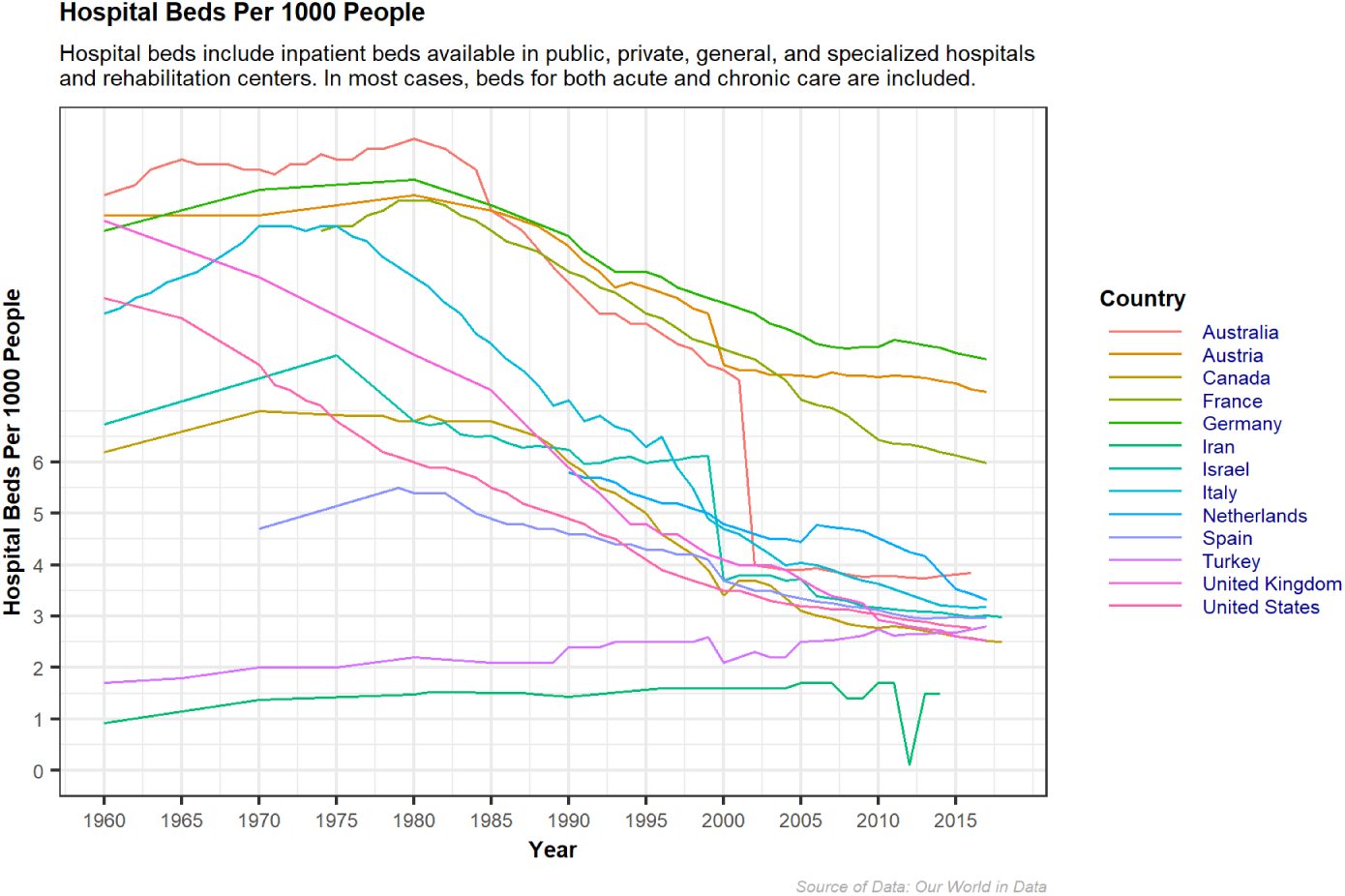
Healthcare Capacity: Hospital Beds per 1000 People.

### 1.3 Crisis analytics: Which metrics should be tracked?

Beside CFR, and crude mortality rate, more metrics were monitored and tracked on a daily basis by the public before the professionals. As mentioned above, each metric reflects one side of reality and can not indicate it all. Therefore, looking at a set of numbers at the same time is more logical, comprehensive and conclusive [10]. The metrics which were reported daily are split into two main categories; *absolute* and *relative* categories. While the *absolute* category included metrics such as: number of confirmed cases, number of deaths, number of recovered cases, number of critical cases and total number of tests; the *relative* category included other metrics such as: death rate (CFR), recovered rate, number of deaths per million people and number of tests per million people. It was a learning curve development exercise for everyone where a mix of different metrics from each category was selected and tracked in an effort to conclude, speculate or even predict the end of the outbreak. It was even obvious that many professionals, media channels reporters and hosts were constructively competing in showing different perspectives based on different sets of metrics. Another critical metric that is essential in the field of epidemiology is the The reproduction number, R0, which is a way of rating a disease’s ability to spread, [11]

Few of them made the effort to create or propose a new metric, or set of metrics, out of the already existing data which might have explained the unusual variations in the behavior of the ongoing COVID-19 pandemic [12]. This resulted in similar conclusions with slight differences that did not provide the decision makers with deeper insights about the pandemic nor with valuable recommendations in dealing with the unpredictable behavior of the outbreak, promptly.

It is a critical step to set an objective, or set of objectives, before deciding on which metrics should be observed and reported. Different objectives require different metrics and numbers to be gathered and analyzed to reach valuable and conclusive findings. In this paper the objective was to assess the effectiveness of the measures and the interventions which took place by the decision makers during the ongoing COVID-19 outbreak. Besides the above objective, assessing the efficiency of the health system in dealing with the outbreak was another important objective in this study to help decision makers react to the unpredictable dynamics of the crisis.

Decision makers in each of the 210 affected countries and territories around the world [13], were in a desperate need to have access to different indicators and metrics that helped them quantify the impact of their interventions. In this paper a new metric was proposed based on the available daily data for both the death rate (CFR) and recovered rate. The detailed method is explained in section 3.

## 2 Crisis is a non-stationary dynamic system

A stationary behavior of a system or a process is characterized by non-changing statistical properties over time such as the mean, variance and autocorrelation. On the other side, a non-stationary behavior is characterized by a continuous change of statistical properties over time [14]. Without the need of a mathematical proof, it was obvious that the COVID-19 pandemic was one of these non-stationary systems, where its behavior was unpredictable and in many situations inexplicable. Dealing with this kind of systems is a complicated task for many researchers and professionals. Nevertheless, during the past few years, scientists and researchers managed to study and introduce various methods and techniques, mainly mathematical models, in attempts to understand and predict the behavior of the non-stationary systems.

The objective of this paper is not to introduce another method, but to theoretically explain the behavior of the dynamic system during an outbreak of a virus like SARS-CoV-2. In this paper, a systems thinking approach was adopted to develop a conceptual model for the COVID-19 pandemic, in an attempt to identify the potential intervention points in the system for decision makers to control the outbreak. Moreover, the systems thinking model allows decision makers to identify which metric, or set of metrics, are more helpful in assessing the effectiveness of their interventions during an ongoing outbreaks.

### 2.1 Systems thinking model for the COVID-19 outbreak

The main purpose of the systems thinking approach [15], is to develop a holistic view of the system under study without the need to build detail models. As the main objective of the paper is to shed light on how the proposed *snr* index (see sections 3 and 4) is used in helping decision makers to see the bigger picture rather than its detailed complexity.

A systems thinking approach is used to explain the dynamism of a system that consists of various components that are interconnected among each other in a cause-and-effect manner forming a coherent structure of multiple feedback loops whose behaviors over time show a positive (reinforcing) and/or a negative (balancing) dynamic, [16, 17]. Moreover, the delay components between the cause and effect factors are articulated explicitly in the model to emphasize their contributions to the dynamism behavior of the system [18, 19, 20, 21].

Figure 4 shows the simplified systems thinking model for the COVID-19 outbreak. The model shows three dominant feedback loops that drive the dynamics of the system during the COVID-19 pandemic. The positive (reinforcing) feedback loop (FBL#0) is being triggered by the outbreak of the virus SARS-CoV-2 in China by the end of December 2019 showing an exponential growth of total positive cases [22]. Nevertheless, for the purpose of this paper, which is to quantify the effect of interventions by different governments, the total confirmed cases, *TCC* (positive diagnosed cases), are considered as the state of the system due to the unavailability of the data for the untested (undiagnosed) positive cases.

**Figure 4:**
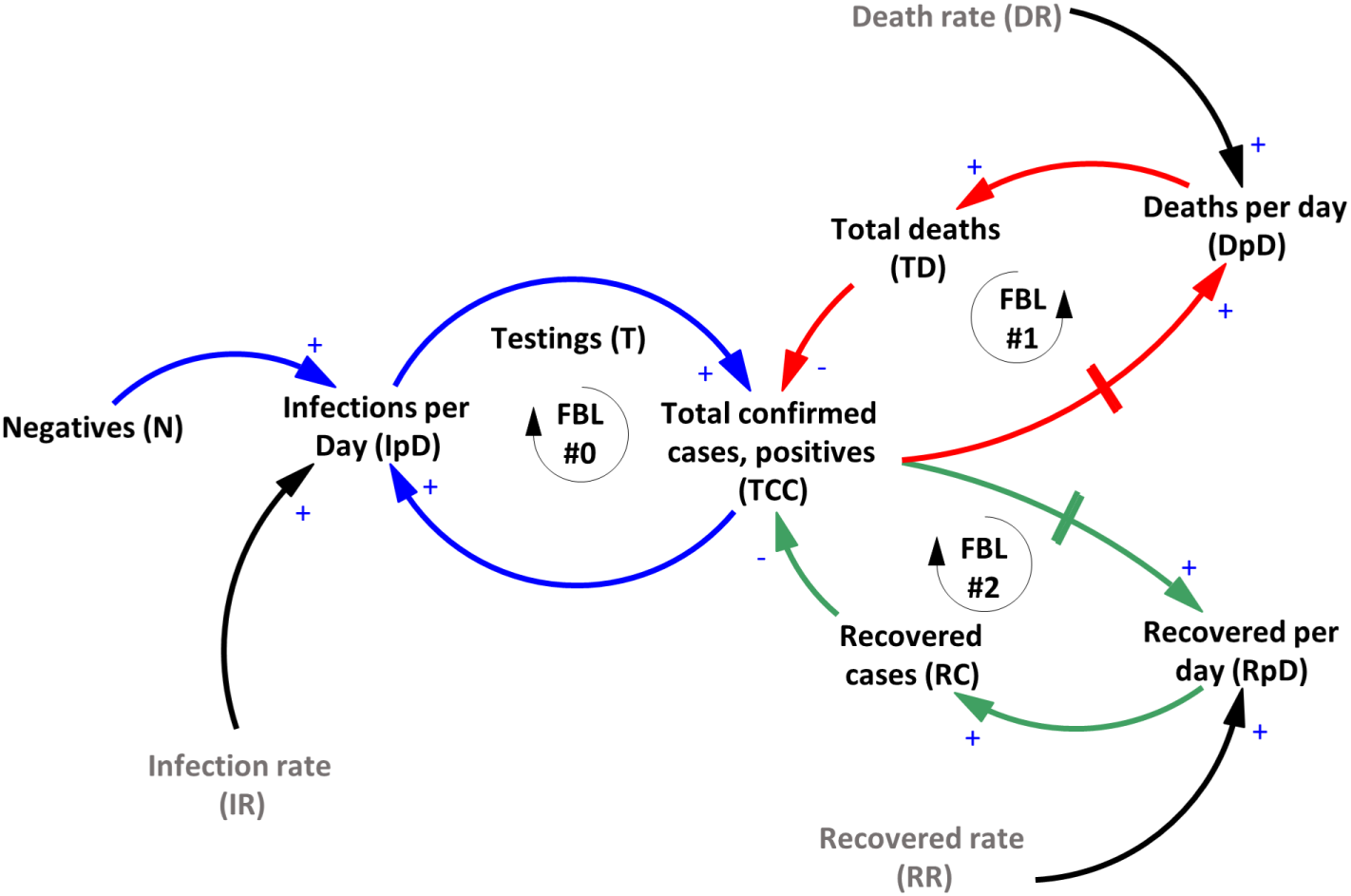
A Simplified Systems Thinking Model for COVID-19 Outbreak with the Main Three Feedback Loops.

As in any exponential growth behavior, the number of infected people (undiagnosed) keeps growing inconspicuously at the early stage of the outbreak driven by the infection rate, IR as shown in the model, figure 4. Without proper testing in terms of timing and volume, the behavior of the dynamic system is inexplicable and uncontrollable. The main feature of any pandemic is that the infection rate, *IR* keeps rising in an increasing rate as long as no governmental responses took place, leaving no choice for the contact rate among people but reaching its highest level which accordingly helps in spreading the virus in an exponential behavior. FBL#0 (infectious loop) shows a vicious circle behavior that is responsible for the outbreak of COVID-19. The aggravated growth of the total confirmed cases is balanced and offset by the behavior of another two dominant and negative feedback loops, FBL#1 (death loop) and FBL#2 (recovery loop), as shown in figure 4.

Both feedback loops, FBL#1 and FB#2 show a typical negative (balancing) dynamic behavior that balances the growth of the total cases (confirmed and unconfirmed). As indicated above, due to the unavailability of the data for the positive cases (undiagnosed), only the total confirmed (diagnosed) cases are considered in this paper.

Irrespective of its pace, the negative (balancing) feedback loop FBL#1 through its rate, *DR* (or *CFR)* adds daily new deaths, *DpD*, which increases the total deaths, *TD*. While the feedback loop FBL#2 through its rate, *RR* adds daily new recovered cases, *RpD*, which increases the total recovered cases, *RC*, as shown in figure 4. Obviously, both feedback loops, FBL#1 and FBL#2 decrease the total number of infected cases, *TCC* in different yet opposite ways. Both feedback loops are competing against each other in terms of speed. The faster the feedback loop is, the higher the resulting corresponding numbers it produces, *TD* and *RC*.

### 2.2 COVID-19 outbreak complete systems thinking model

For the purpose of completeness of the model, figure 5 shows the complete systems thinking model including a positive polarity feedback link from *RC* to the negative cases (not infected cases), *N*, resulting in an increase of the healthy people. Moreover, to calculate the death rate, *DR* and the recovery rate, *RR*, a positive and a negative polarity feedback link are added from the total confirmed cases, *TCC*, total deaths, *TD* and recovered cases, *RC* to both *DR* and *RR* respectively, as indicated in figure 5.

**Figure 5:**
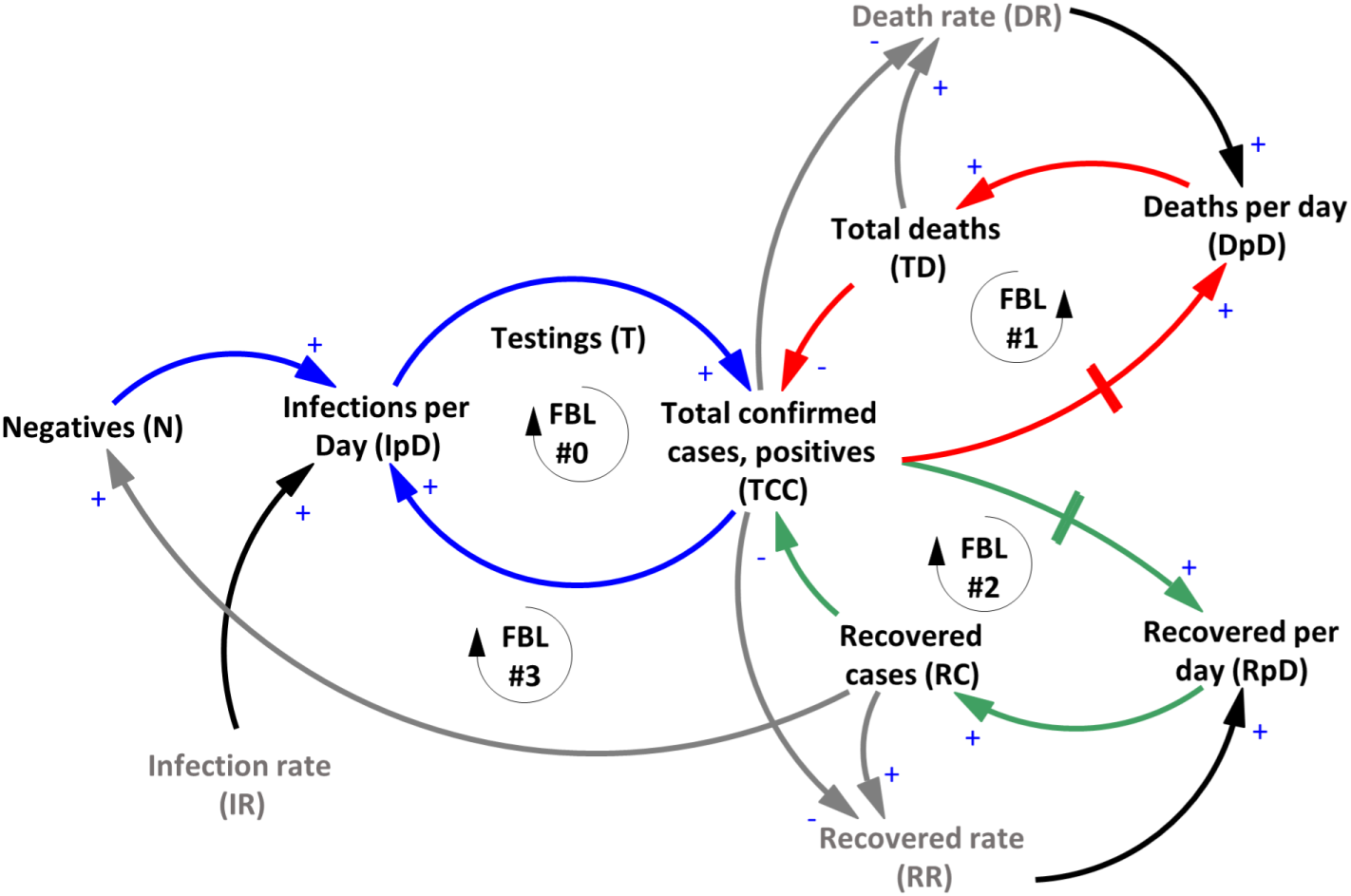
The Complete Systems Thinking Model for COVID-19 Outbreak.

## 3 The Signal-to-Noise ratio: An overview

In the literature of science and engineering, signal-to-noise ratio, *snr* [23], is a common concept and used to compare two levels of a channel or a medium. The two levels are: the desired part of the channel; called signal, *S*, and the unwanted part; called noise, *N*. While *snr* is a common concept in engineering, it has also been applied in different fields and disciplines such as biochemical signaling between cells, or financial trading signals [24, 25].

It usually is defined as the ratio of signal power, *P_S_* to the noise power, *P_N_* [26]. A ratio higher than *1* indicates more signal than noise, which is the desired state. Both signal power, *P_S_* and noise power, *P_N_* must be measured at the same or equivalent points in a system, and within the same system bandwidth (time). Power, *P* is another term that is applied in various fields with different definitions. In physics, power, *P* is the rate of doing work, *W* or of transferring heat, i.e. the amount of energy transferred or converted per unit time. The equation of power can be written as the rate of work:

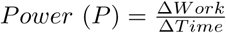

where work, *W* is measured in terms of a net change in the state of the physical system. Accordingly, the *snr* can be calculated as follows,

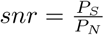

It is important at this point to stress that power, *P* is a quantity that requires a change in both, the physical system and a specified time interval in which the change occurs.

Borrowing the above concepts and applying them to the health system which is dealing with the COVID-19 outbreak, work, *W* represents the net of change in both, the total deaths, *TD*, and the total recovered cases, *RC*. In other words, work, *W* is the effort that the health system, including personals and other capabilities, must carry out to change (transform) the state of the positive cases (inflow of patients) to recovered cases and to prevent or minimize the deaths. Accordingly, power, *P*; in this paper represents the amount of effort/work done by the system during a unit of time (in this paper it is a daily change) to minimize the noise in the system (deaths) and to magnify the signal (recovered cases). Both power terms, *P_S_* and *P_N_*, of the signal-to-noise ratio, *snr* are defined by the recovered rate, *RR* and the death rate, *DR*, respectively.

According to the very definition of the *snr*, the health system efficiency and the effectiveness of the governmental interventions can be assessed at a specific date, *t*, by calculating the logarithm value of the ratio between the recovered rate, *RR*, representing the power of the signal (the wanted part) and the death rate, *DR*, representing the power of the noise in the system (the unwanted part), at a specific day (time, *t*) as follows,

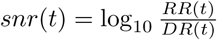

The resulting value (*snr* index) indicates how efficient the system is in minimizing/preventing the deaths and magnifying the recovered cases.

It is important, at this stage of the calculation to clarify that there is no developed benchmark or a scale yet in assessing whether the *snr*, index indicates a good or bad performance of the system under investigation against a known scale. Nevertheless, if the above index is multiplied by a factor of 10, the resultant value would carry a decibel unit (dB). This international standard unit (dB) is used in many disciplines, yet, it is too early to come to a final conclusion whether the resultant values from this method can be compared to other dynamic systems. Accordingly, in this paper, comparative scenarios were developed among different countries without paying attention to the meaning of the unit as explained above.

In this paper, a decibel (dB) unit is used to help in contrasting the differences among various countries’ performance during the ongoing COVID-19 outbreak. As a result, the following equation is proposed to calculate the *snr* index:

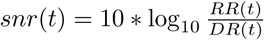

In all cases, an index, *snr* value greater than 0 (positive values) indicates clearly that the system under investigation is performing better than a system with an index value smaller than 0 (negative values). Moreover, a final value of the *snr_f_* index can be established by the following equation given the value of the death rate, *DR* at time, *t* and assuming that all other active cases will be recovered at the end of the crisis:

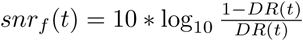

or

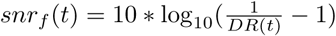

Using this *snr_f_* value, the decision makers could set a target for the *snr* index during the crisis.

## 4 The *snr* index in practice: Assessing the intervention’ effectiveness and system’s efficiency

As discussed in section 1.3, reporting the rate of each feedback loops (see section 2.1) separately will neither help the stakeholders take the correct intervention at the proper time nor assess the system’s efficiency and the effectiveness of their interventions.

### 4.1 Intervention points and types

Figure 6, shows possible three points of interventions to the system, namely, at the infection rate input, *IR*, at the death rate input, *DR* and at the recovered rate input, *RR*. The three intervention points were developed to evoke two main impacts, (1) slowing down the spread through the lockdown, social (physical) distancing, etc. (see section 1.1), and (2) fixing the problem through, (a) stopping the death feedback loop #2, and (b) accelerating the recovery feedback loop #2. Fixing the problem interventions required the health system to be ready and equipped with the needed testing kits and facilities, beds, equipment, workers, etc. to manage the crisis by managing the dominant feedback loops as discussed above.

**Figure 6:**
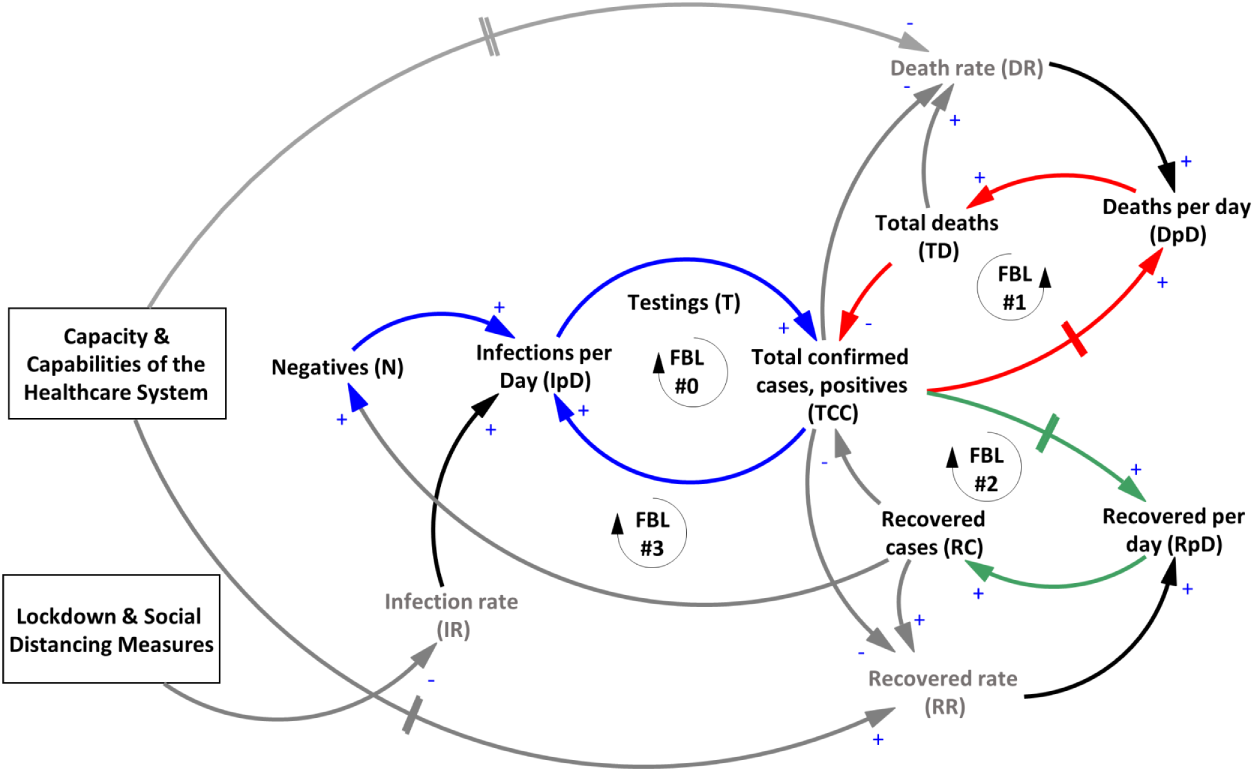
The Three Main Intervention Points in the COVID-19 Dynamic Model.

### 4.2 Assessing the system’s performance through the *snr* index

In this study the *snr* index was proposed, as discussed in the previous sections, to answer the above mentioned questions regarding systems’ efficiency and interventions’ effectiveness. Figure 7 shows the complete systems thinking model including the three intervention points and the way, the *snr* index is calculated.

**Figure 7:**
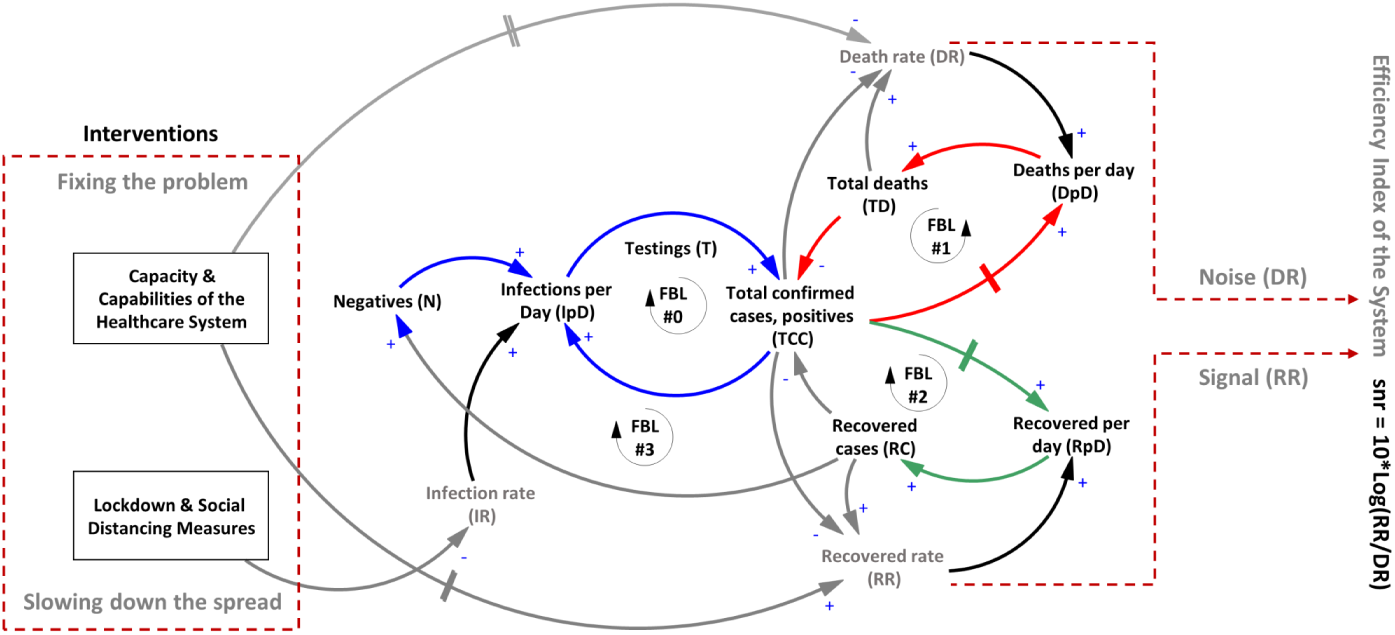
COVID-19 Dynamic Model Showing the Calculation of the *snr* Index.

It is important, at this stage, to mention that although this *snr* ratio seems to produce the same value as when the total recovered cases are divided by the total deaths on a specific day, nevertheless the interpretation is different, as the later does not take into consideration the pace (rate) of the feedback loops. As for the former ratio, *snr* index, the signal refers to the recovered cases, representing the gain in the system, which is the desired situation. On the other side, the noise component of the *snr* index refers to the number of deaths representing the losses in the system. A higher value of the *snr* index greater than *0* indicates one of three scenarios: (see figures 4 and 5) (1) the pace (rate) of feedback loop FBL#1 (death loop), that is controlled by the death rate *DR*, is slowing down while the feedback loop FBL#2 (recovery loop) stays relatively constant, resulting in a slightly increase in the total deaths, (2) feedback loop FBL#2 (recovery loop), that is controlled by the recovered rate *RR*, is speeding up while feedback loop FBL#1 (death loop) does not change much, resulting in a slightly gain in the system by decreasing the positive cases (infected) and hence increase in the recovered cases, or (3) feedback loop FBL#2 (recovery loop) is accelerating while feedback loop FBL#1 (death loop) is decelerating which is the desired scenario for an efficient health system and high effectiveness of the interventions. If this last scenario takes over the situation, the *snr* index will increase until it theoretically and mathematically reaches an undefined value marking the end of the pandemic. This situation is dominant during the pre- and post-crisis, therefore this proposed *snr* index method is feasible during the crisis.

Using this proposed metric *snr*, it is possible to compare different countries’ health systems in terms of efficiency and effectiveness of the interventions during the ongoing crisis. Since, the *snr* index is a logarithmic value as discussed in the previous section, a higher positive value of the *snr* index indicates high efficiency of the health system that is able to speed up the testing process, and the recovery RR, while keeping the death rate *DR* at its slowest level. On the other hand, a value of *snr* index closer to zero, indicates both rates; *RR* and *DR* are almost equal, which indicates that most of the carried out interventions are not working as anticipated. Obviously, a negative value of the *snr* index indicates that the system is extremely inefficient and most of the interventions are not working indicating a chaotic behavior in managing the crisis, which might lead to a system’s failure and collapse.

In the next section, the interventions’ effectiveness and system’s efficiency during the outbreak of different countries will be discussed in the light of the proposed *snr* index.

### 4.3 Selection criteria for countries comparison

In order to reach plausible findings, a set of criteria had to be defined to select the countries for comparison purposes. According to the paper’s objective, only the countries where their systems were highly strained during the outbreak, were selected. To reach these criteria, a 2-dimensional graph showing the relationship between the total confirmed cases and the confirmed cases per 1 million people was constructed for a subset of countries, see figure 8. Accordingly, countries that are in the upper right quadrant of figure 8 were selected for further analysis. These countries have more than 5,000 confirmed cases and more than 200 confirmed cases per million people. Countries that had a relatively small number of confirmed cases were excluded from the selection, because in these countries, the death rate, *DR* and the recovered rate, *RR* are particularly poor metrics and not indicative of the real situations due to the lack of proper and frequent testings as well as reporting mechanisms.

**Figure 8:**
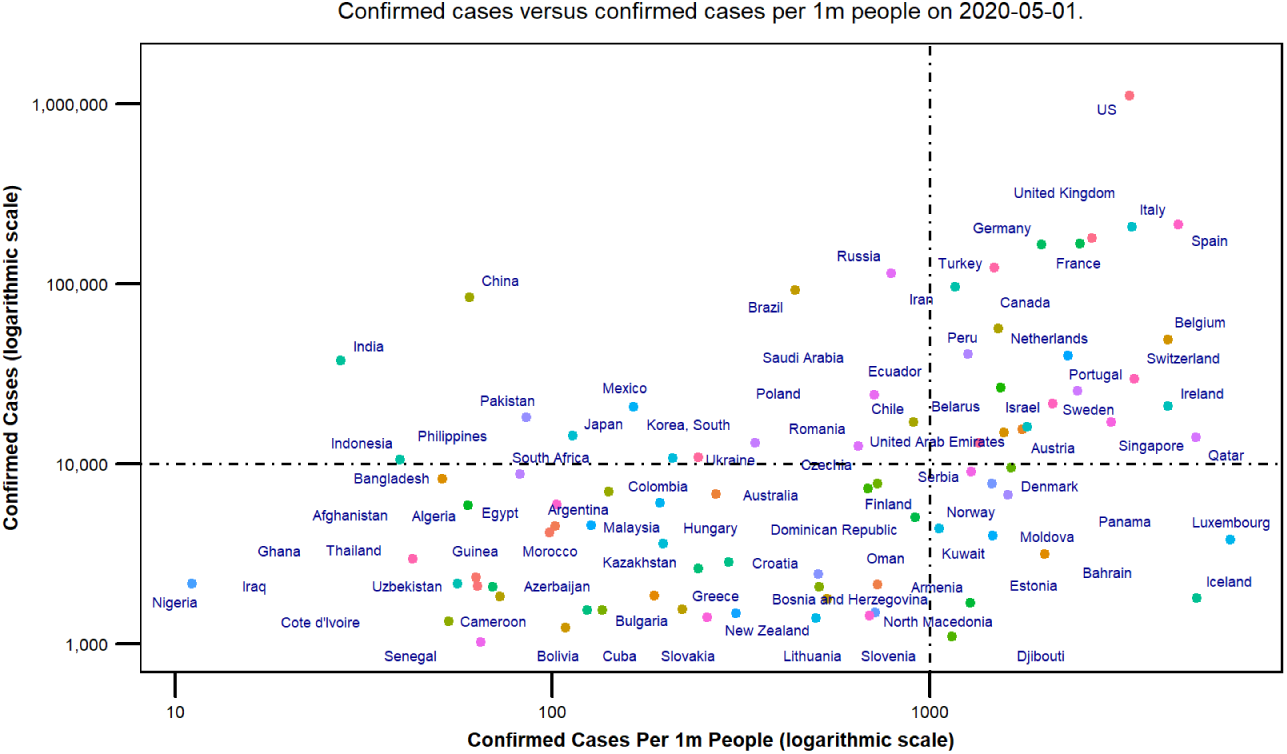
The total confirmed cases versus total confirmed cases per 1 million people for a subset of countries.

Out of the 212 affected countries and territories by the SARS-CoV-2 virus, only 19 countries were selected for comparison purposes. It is important to mention that this number of countries changed daily as a new dataset was updated. Moreover, the selected countries were classified into different groups according to their *snr* scores and behaviors during the outbreak, see figure 11.

Figures 9 and 10 show bar charts for all selected countries according to the criteria defined above, indicating the confirmed cases as well as the death rate, *DR* at the specified date. It is clear from the figures that the USA has the highest confirmed cases due to the massive number of testings they have carried out. Moreover, Spain has the highest confirmed cases per 1 million people due to its’ population size.

**Figure 9:**
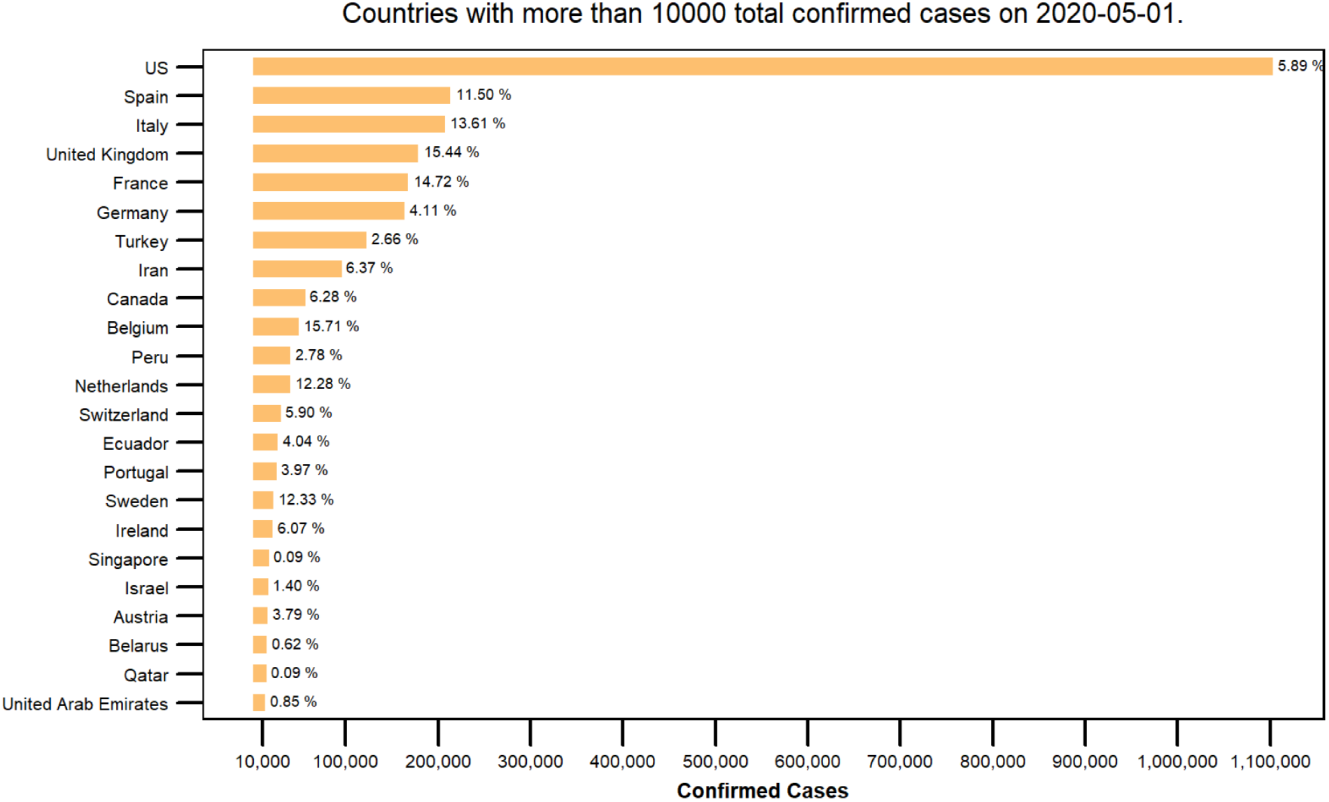
The Confirmed Cases and the Death Rate of the Selected Countries.

**Figure 10:**
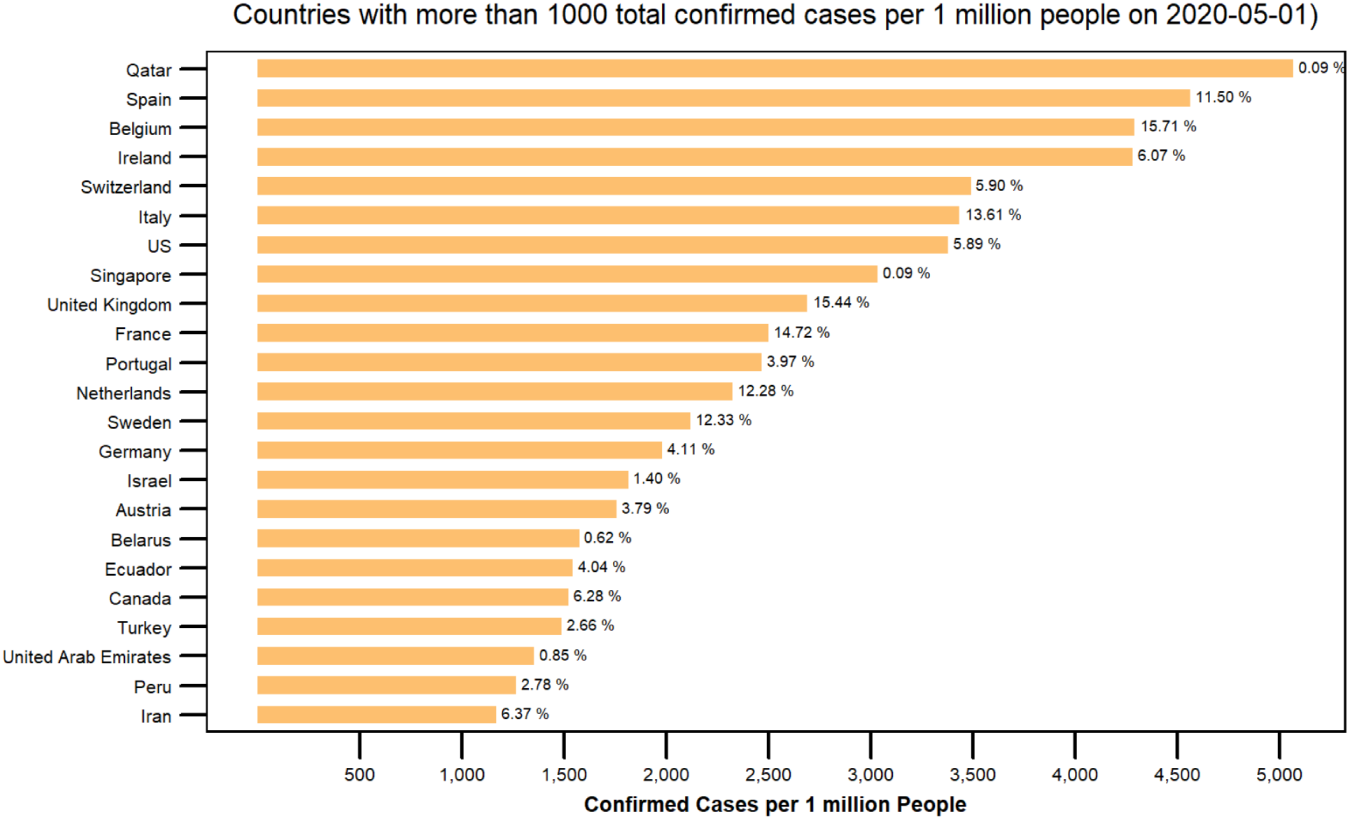
The Confirmed Cases per 1 Million People and the Death Date for the Selected Countries.

Figure 11 gives an overview about the position of each country in a *Death Rate-Recovered Rate* matrix with respect to the world death rate, *DR* (horizontal line) and the world recovered rate, *RR* (vertical line) at the specified date. The figure shows clearly that Germany, Austria, and Switzerland are scoring relatively low on the death rate measure and relatively high on the recovered rate scale, indicating a better position than the rest of the countries. Therefore, these countries labeled as the *benchmark group* which will be used as a reference during comparison with other groups. On the other side, the matrix shows three other groups of countries which either had been *struggling*, *less struggling* or *extremely struggling* during the COVID-19 outbreak.

**Figure 11:**
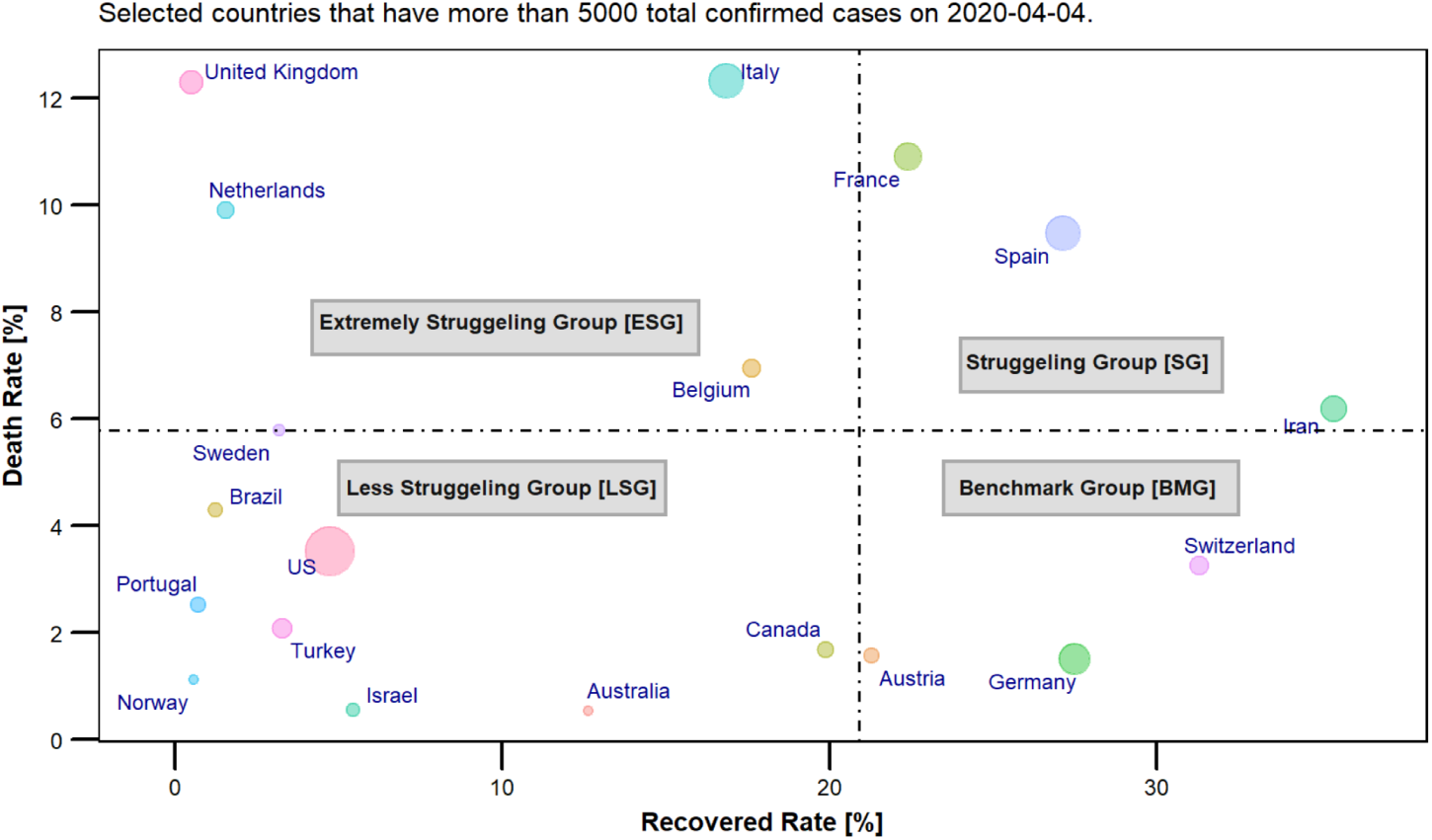
The Death Rate - Recovered Rate Matrix.

In the next sections, different scenarios were developed to compare the countries’ performance (the *snr* curve) during COVID-19 outbreak starting 1st of March 2020 to the specified date.

### 4.4 The *snr* curve for the benchmark group

In this section, the *snr* curve will be discussed showing how each country was reacting to the spread of the SARS-CoV-2 virus. As shown in figure 12, the *snr* curve starts for each country at a higher level (until first death case) of the *snr* index and goes down very fast to a lower level at the early stage of the outbreak. In this group, although the outbreak started earlier for Switzerland than for the other members in this group, it a faced very hard time due to the turbulence created by the spread of the virus. Its *snr* index even went to the negative zone of the index between 10*^th^* of March till 24*^th^* of March. Afterward, it managed to control the situation and ever since it has been going up to a relatively high *snr* index reaching a level of 20 dB around the 2*^nd^* of April and maintaining its stabilized position at the later stage of the outbreak. Comparing Switzerland’s *snr* behavior against the other two members of the benchmark group, Germany shows by far an extremely strong performance during the outbreak where its *snr* curve went down slightly beneath the level of 10 dB for a very short time (3-4 days), yet within the positive zone of the *snr* index. Indubitably, Germany was the most prepared country in the whole world in facing such pandemic with a strong health system (see section 1.2) and well planned interventions on all levels, time-wise and clarity. Germany’s *snr* curve reached its highest level at around 30 dB by the 23*^rd^* of March marking the shortest period of time that any country had to go through to stabilize its system. Austria on the other side, started the fight against the pandemic later than Switzerland and Germany, but nevertheless managed to reach the positive zone of the *snr* index around the 25*^th^* of March and even outperformed Switzerland by the 28*^th^* of March.

**Figure 12:**
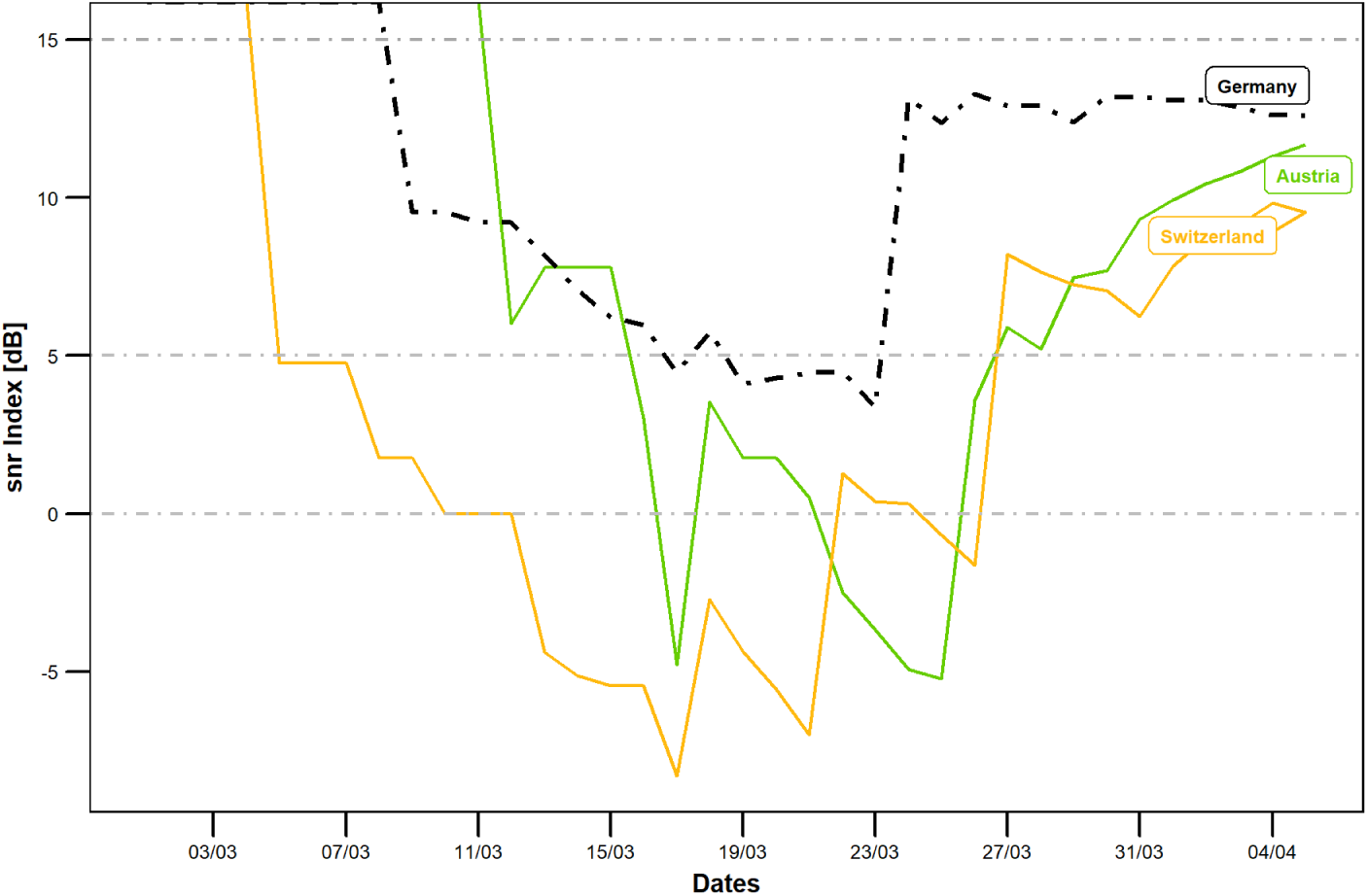
The *snr* Curve for the Benchmark Group Countries, starting 1*^st^* of March 2020.

The *snr* curve for the *benchmark group* follows an interesting pattern (square root shape like) passing through four main episodes, see figure 13. The starting episode, *episode 0* (downward), where the system is being shocked by the outbreak of the virus taking the whole system into a deep and steep dive to a lower level of the index that usually reaches the negative zone except for Germany in this group. Episode 0 is characterized by an adhoc and rather delayed intervention from the stakeholders, who were trying to figure out the situation and actions to be taken. The second episode, *episode I* (unstabilized horizontal), is where the stakeholders had to take more drastic measures to contain the outbreak and to flip the pattern. Many countries, as will be shown in the next sections, could not make it easily due to a lack of testing kits and facilities, delayed interventions, strained healthcare system and more. Countries like the *benchmark group* managed to survive this episode I relatively quickly ranging from 1 to 2 weeks during March 2020. Third episode, *episode II* (upward), is characterized by a rise up of the *snr* curve to a high level of the *snr* index reaching a value between 25 and 30 dB for this group. Once the curve reached its highest level, the fourth episode III starts marking higher stability in the system for probably a while till this cycle repeats itself, should the second wave of the outbreak start again, otherwise the country can start its plan to gradually relax the restrictions.

**Figure 13:**
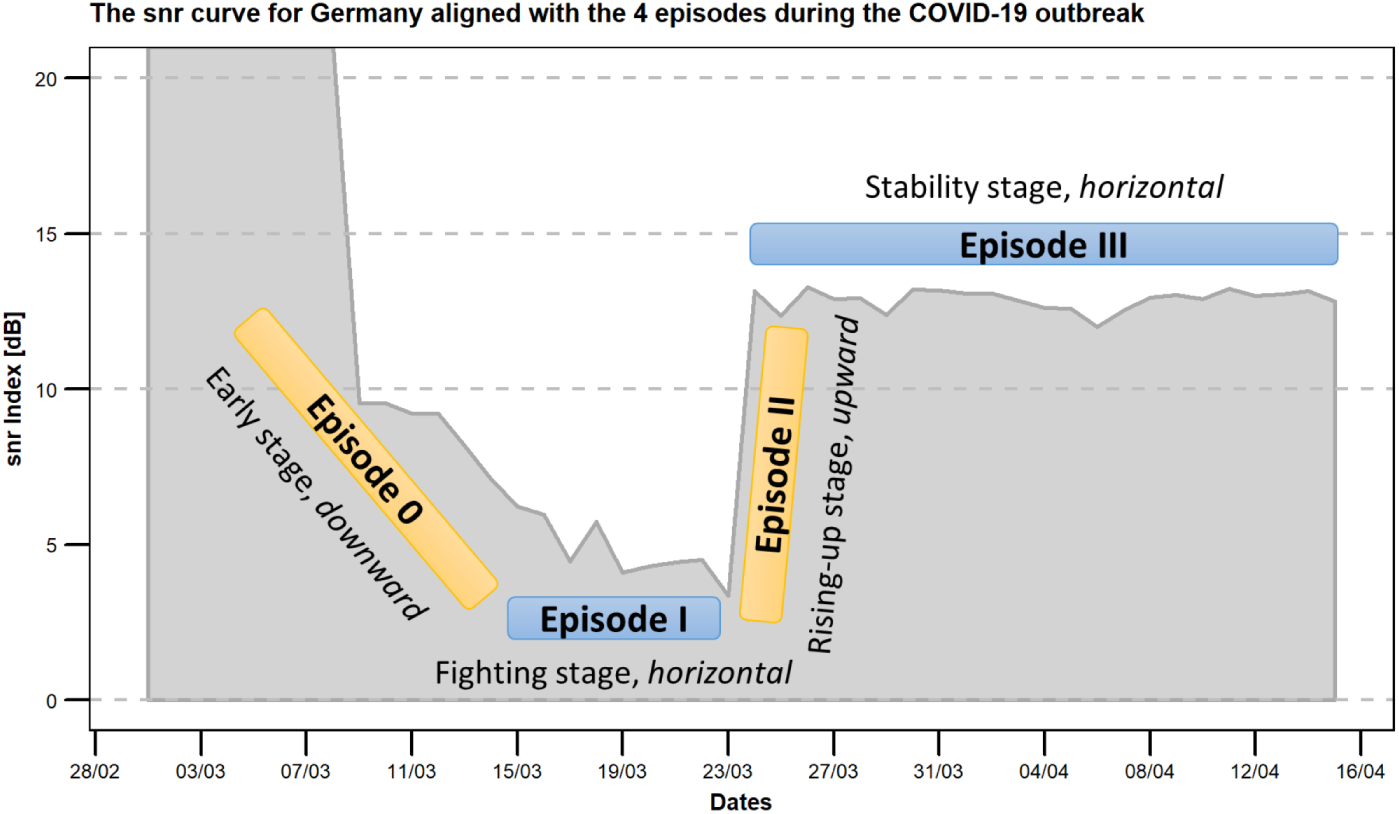
The *snr* Curve for Germany aligned with the 4 Episode Pattern, starting 1*^st^* of March 2020.

Before discussing the *snr* curve for the other groups, it is worth comparing the behavior of the *snr* curve of the benchmark group against South Korea’s. Although, South Korea, as per figure 14, shows extremely high performance, it was not selected among the countries which fall within the predefined criteria, see section 4.3. Another reason for why S. Korea was not shown in further analyses is that the data available for South Korea in the dataset used for the analysis did not include the period of time where the country started the fight against the outbreak. Moreover, S. Korea, based on its past experience with other outbreaks such as MERS, its health ecosystem, data privacy legality issues and technological capabilities were ready and clear, which would make the comparison biased and invalid.

**Figure 14:**
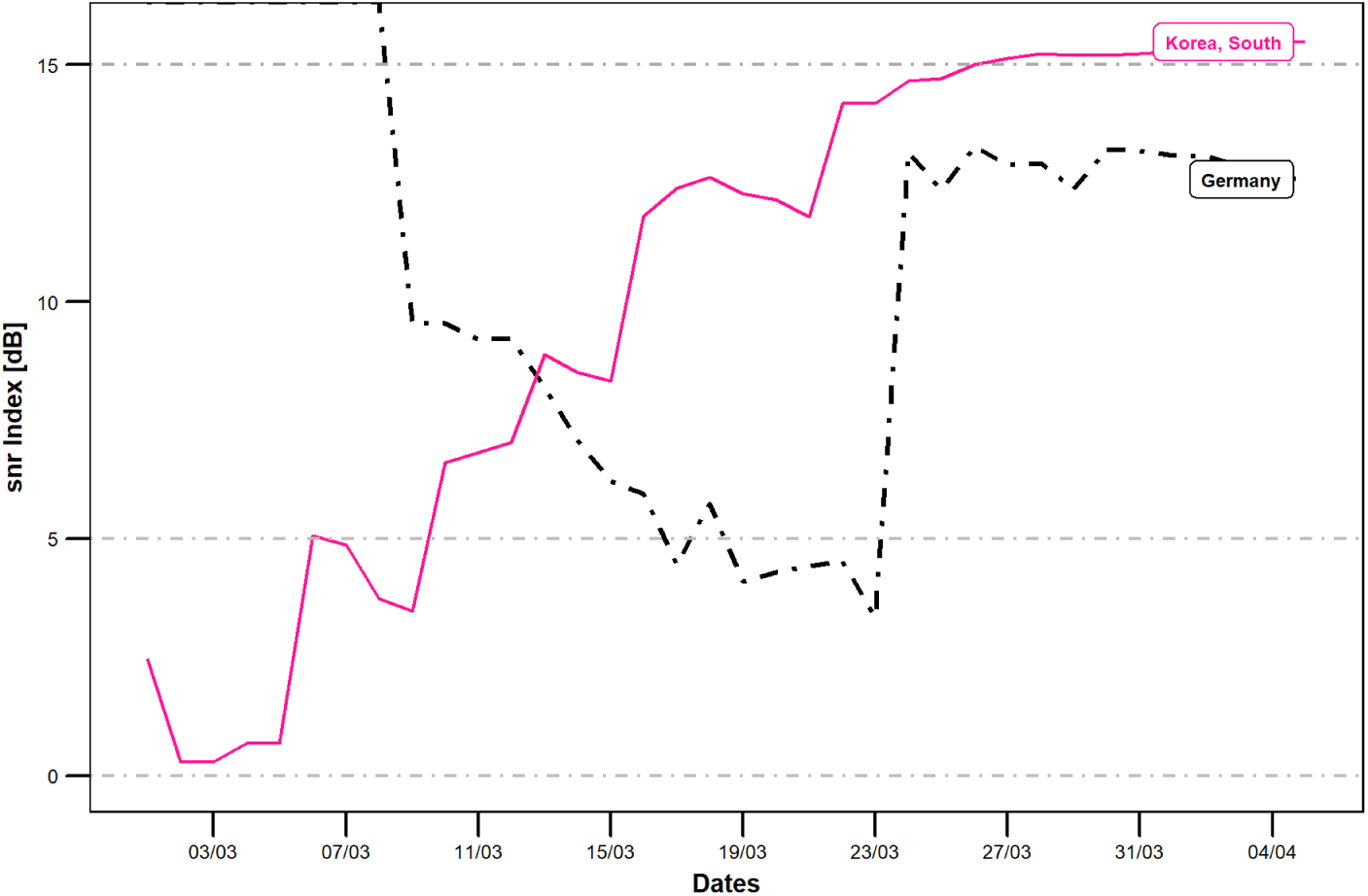
A Comparison of the *snr* Curve between South Korea and Germany, starting 1*^st^* of March 2020.

In the next section, various countries from the other predefined groups will be compared to Germany only and not against the whole benchmark group in order not to populate and hence distract the data visualization.

### 4.5 The *snr* curve for extremely struggling group compared to Germany

As mentioned before, and shown in figure 11, this group had the highest death rate, *DR* and relatively the lowest recovery rate, *RR*, among the other selected countries for this study. This group included France, Italy, the United Kingdom and the Netherlands. In figure 15, the *snr* curve of the above mentioned countries show how the situation was deteriorating for them at the early stage (episode 0) of their fights against the outbreak. Although Italy was on the daily news around the globe showing worries about the country and its people, surprisingly its *snr* curve shows it has been doing relatively well where the *snr* curve never crossed the zero level to the negative zone as with the other three countries in this group. In this section Italy’s case will not be discussed but will be shifted to section 4.8 where other countries which showed an irregular *snr* curve behavior and will be discussed separately.

**Figure 15:**
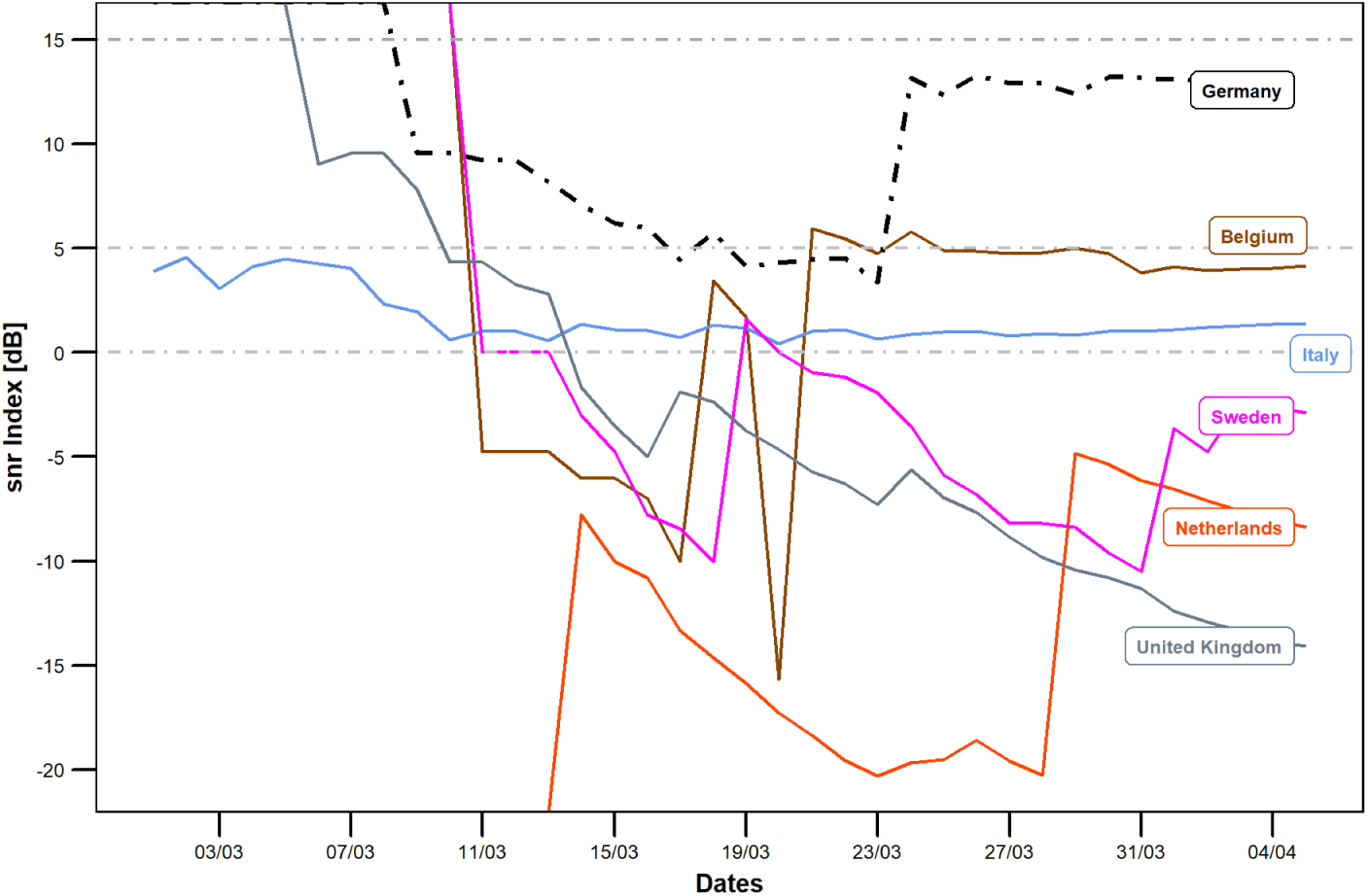
The *snr* Curve for the Extremely Struggling Group Countries Compared to Germany, starting 1*^st^* of March 2020.

Episode 0 (see section 4.4 for more details) of the *snr* curve for the three countries, France, the UK and the Netherlands shows a deep dive into the negative zone of the *snr* index indicating that neither the early interventions taken by the decision makers were working nor was the health system capable of coping with the pace of the outbreak. While France managed to flip the *snr* curve during 21-22 March, the UK and the Netherlands were still struggling and stayed in the negative zone at the time of this study. Figure 15 shows that the Netherlands seemed to find some working interventions for a while but that they were not enough to lead the *snr* curve up to the positive zone and rather stayed in the negative zone at a little higher level after 29*^th^* March. It is worth mentioning at this point that the *snr* curve for the Netherlands completed one full cycle (episode 0 to I then to II and III) in the negative zone demonstrating that the Netherlands requires another cycle of the curve to reach the positive zone. The UK on the other side was not able at all to show any progress during episode 0 and never reached episode I, indicating clear failure on both sides; the interventions and the healthcare systems.

### 4.6 The *snr* curve for struggling group compared to Germany

Figure 16 shows the *snr* curve for a group of countries whose metrics marked relatively high values on both axes, death rate, *DR* and recovered rate, *RD*. Except Iran (see section 4.8 for more discussions about irregular *snr* behavior), the *snr* curve for the other two countries in this group, Spain and Belgium showed clearly the expected behavior; four episodes, 0, I, II and III (refer to section 4.4, for detailed explanation). From the figure, it was obvious that Spain found its way to a stability (positive zone) around 23*^rd^* of March after some considerable fights against the outbreak that started around 4*^th^* of March. Moreover, its *snr* curve shows multiple shorter cycles during the above mentioned period ended by 23*^rd^* of March. During this period, the decision makers were able to put effective interventions in place supported by the relatively well coping healthcare system (see section 1.2 as well as figures 2 and 3), despite Spain showing the highest confirmed cases in one million people among the selected countries for this study (see figure 10), indicating high strain on its healthcare system during the pandemic.

**Figure 16:**
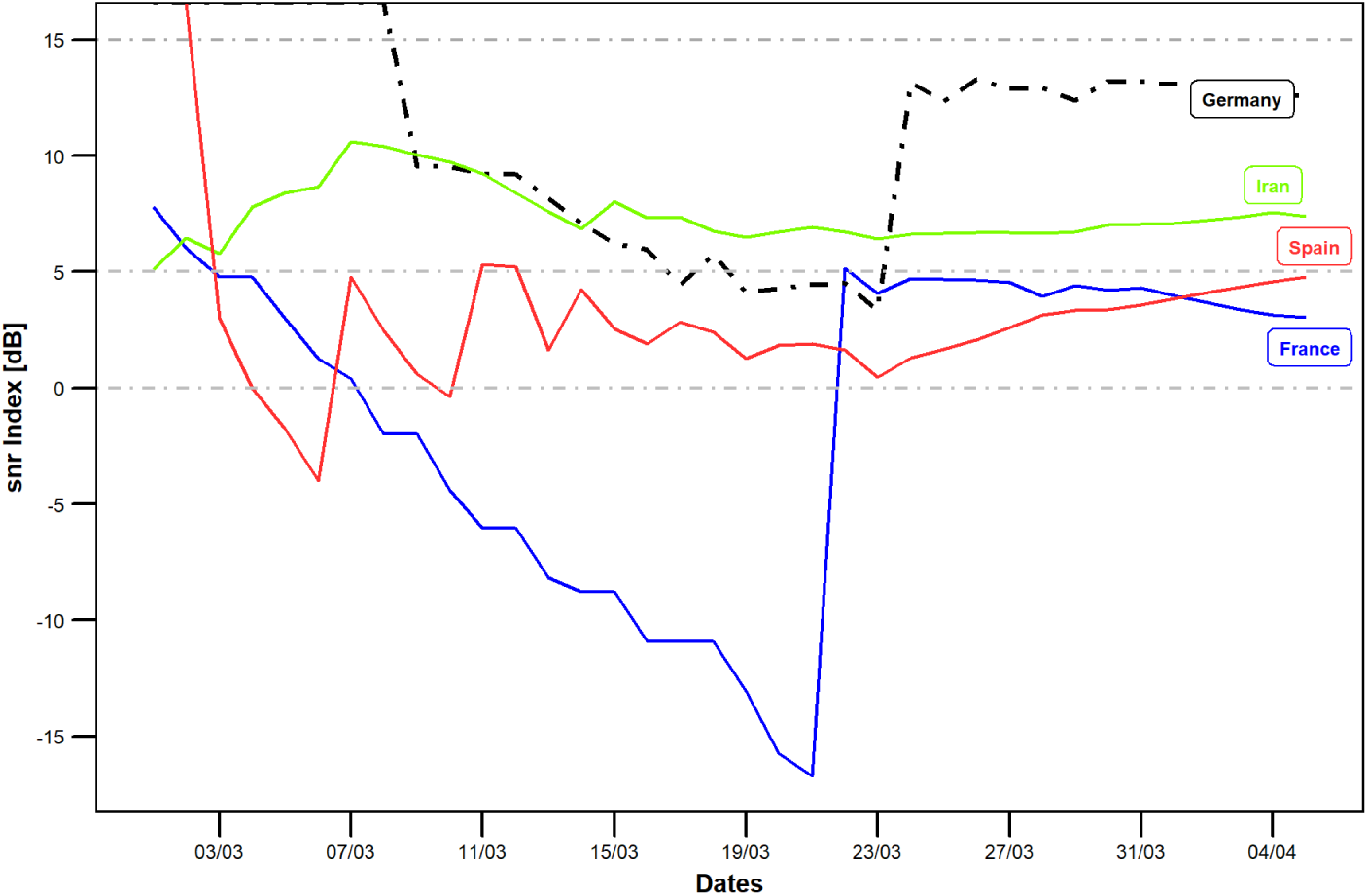
The *snr* Curve for the Struggling Group Countries Compared to Germany, starting 1*^st^* of March 2020.

Belgium on the other side, had to face a hard fight against the COVID-19 outbreak at its early stage (see the negative zone, between 11*^th^* and 21*^st^* of March), until it managed to enter the positive zone of the *snr* curve by 21*^st^* of March and maintain its position ever since.

### 4.7 The *snr* curve for less struggling group compared to Germany

In comparison to Germany, countries in this group included Canada, the USA, Turkey, and Israel which showed more or less similar behavior with regards to the *snr* curve, see figure 17. Their behaviors followed the same four episode structure of the *snr* curve where episode 0 was mostly spent in the negative zone except for Israel which showed the least death rate, *DR* among the 20 selected countries in this study. This could be attributed to a relatively lower strain on its health system as per figures 2 and 3 as well as 10. Canada’s *snr* curve on the other side shows some similarity to Germany’s curve except the duration between 19-23 of March, where the curve (episode 0) slipped slightly into the negative zone of the *snr* index indicating some struggling of the healthcare system during this period. Although Turkey entered the fight against the pandemic later than many other countries starting the 24*^th^* of March, it surprisingly managed relatively quick within one week, to enter the positive zone of the *snr* curve. As for the USA, it was extremely hard work for all stakeholders trying to bring stability to the system during the early stage of the outbreak. Nevertheless, USA could not enter the positive zone of the curve to follow its members in the group before the 29*^th^* of March marking the longest episode I for this group and hence indicating the least measures with regards to the healthcare system readiness and efficiency. On the other side, comparing all group members including Germany, the USA was at the lowest positive *snr* index level around mid of April 2020.

**Figure 17:**
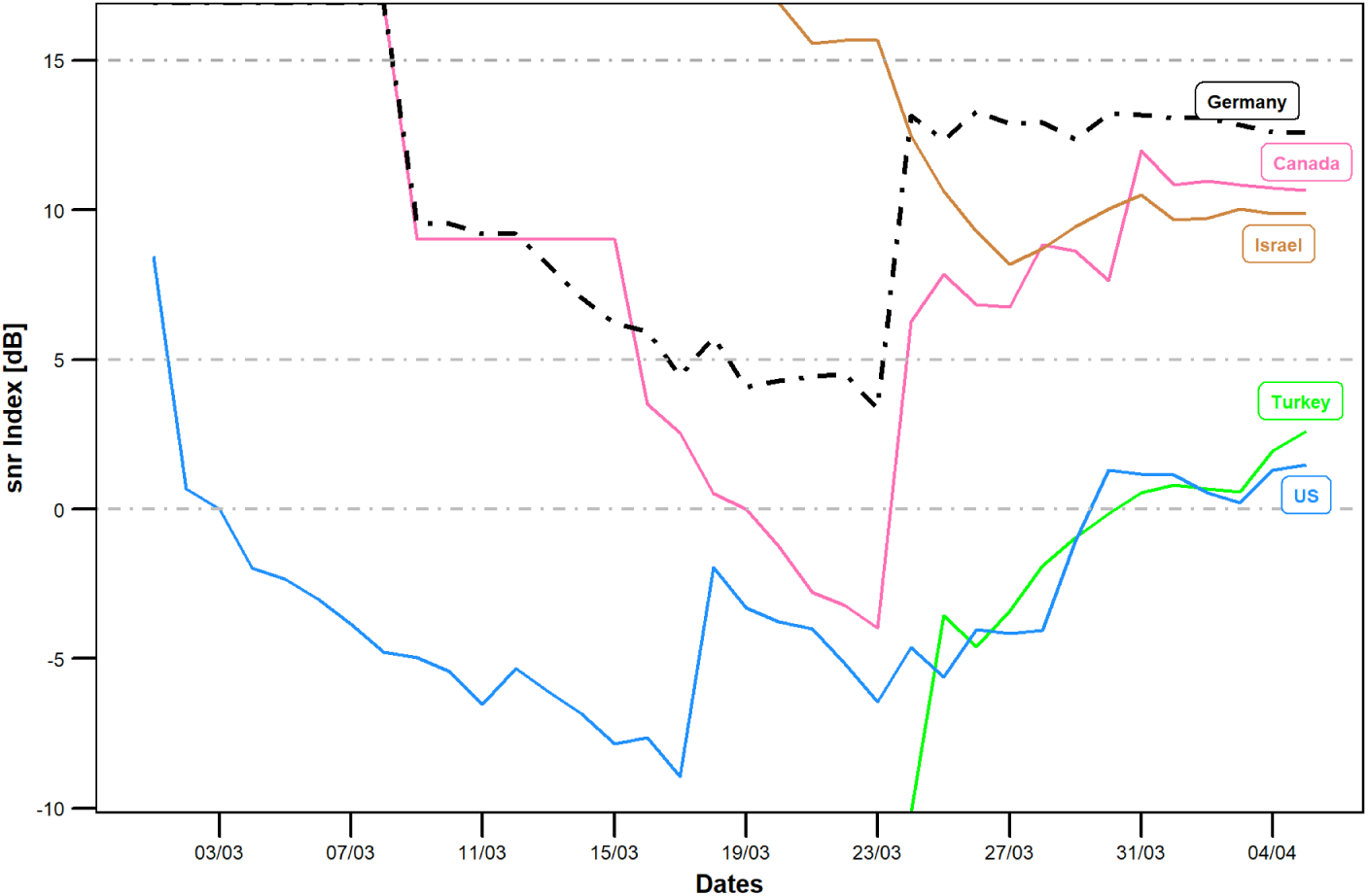
The *snr* Curve for the Less Struggling Group Countries Compared to Germany, starting 1*^st^* of March 2020.

### 4.8 The *snr* curve for irregular group compared to Germany

It was imperative to look further to the *snr* curve’s structure for some countries whose behavior did not match the usual *snr* pattern as is the case with the other countries. Figure 18 shows the behavior of the irregular group members in comparison with Germany’s. As discussed before in section 4.4, the *snr* curve structure consists of four consecutive episodes for a single cycle that can repeat itself depending on how each country deals with the pandemic. For this group, the *snr* curve during episode 0 (early stage of the pandemic) is neither as steep, nor reached the negative zone of the *snr* curve as it was with the countries in the other groups (see previous sections) during the same episode. According to the definition of the *snr* curve (see section 3), this should indicate that all these countries did not encounter difficulty in their fight against the outbreak. In reality, this was not the case. At the early stage of the COVID-19 outbreak, Italy and Iran were reporting high numbers of confirmed cases and deaths. This opens a wide discussion regarding the reliability of the reported data from both countries. To stay in the positive zone of the *snr* curve, the recovery rate, *RR* has to outperform the pace of the death rate, *DR* as per the definition of the *snr* index. Both countries shared the same characteristics of the *snr* curve where they spent a relatively shorter time during episode 0, while spending a longer time in episode I than any other country in any group. Accordingly, it can be concluded that both countries had to release some cases as quick as possible to free spots for the critical cases which eventually increased the recovered rate, *RR* and hence resulted in a positive *snr* index. On the other side, the figure shows a relatively similar behavior of the *snr* curve for both Egypt and India. Although both countries did not report any difficulty in their efforts facing the outbreak, neither country managed to come out from episode I till mid April. This kind of conflicting behavior opens once again a discussion around the reliability of the reported data. As for Israel, as mentioned in a previous section, it did not face difficulty in its fight against the pandemic due to its health system readiness and capacity, see section 1.2. The *snr* curve with its four episodes occurred during an extremely short time in the case of Israel marking the shortest *snr* curve cycle among all selected countries for this study. China’s curve clearly is showing a dominant episode III part where stability was reached before all other countries.

**Figure 18:**
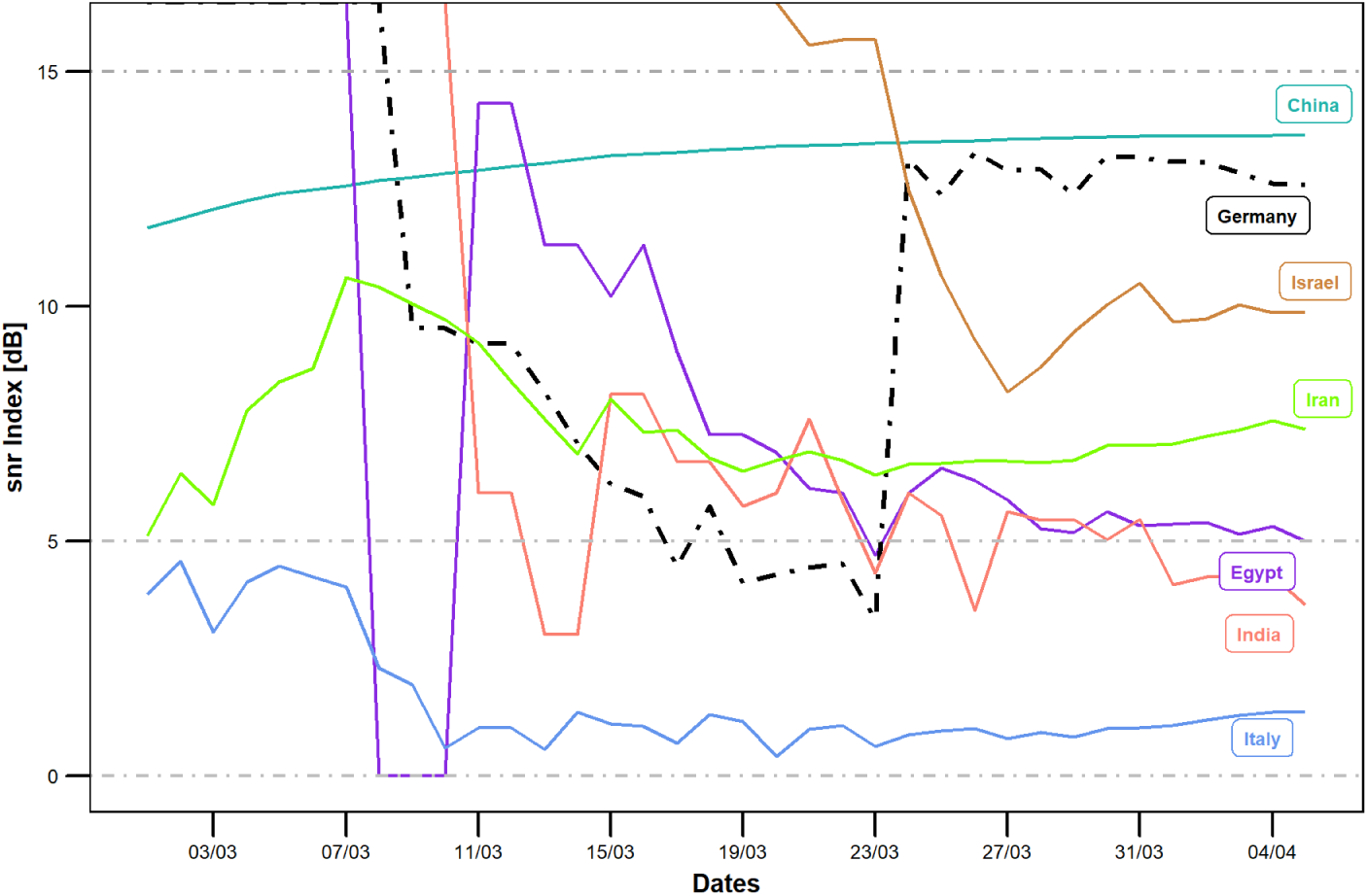
The *snr* Curve for the Irregular Group Countries Compared to Germany, starting 1*^st^* of March 2020.

In conclusion, the countries in this group were showing conflicting results that are not matching with what was happening on ground.

### 4.9 Interventions impact on the *snr* curve: Case of Germany

This section highlights on how the *snr* curve is reflecting the impact of the interventions that took place during the outbreak by the stakeholders. Figure 19, shows the *snr* curve for Germany in association with various intervention types (for more details, see section 1.1) as well as with the ratio between active and confirmed cases. The figure reveals that the interventions such as: movement restriction, public health measures and social distancing took place as early March. With its strong health system in terms of hospital capacity and health workers availability, see section 1.2, Germany was able not only to stay in the positive zone of the *snr* curve during episodes 0 and I, but also to rise up quickly during episode II to reach episode III between 23-24 of March marking the fastest nation to find its stability and maintain its positive position until the writing of this study, see section 4.4.

**Figure 19:**
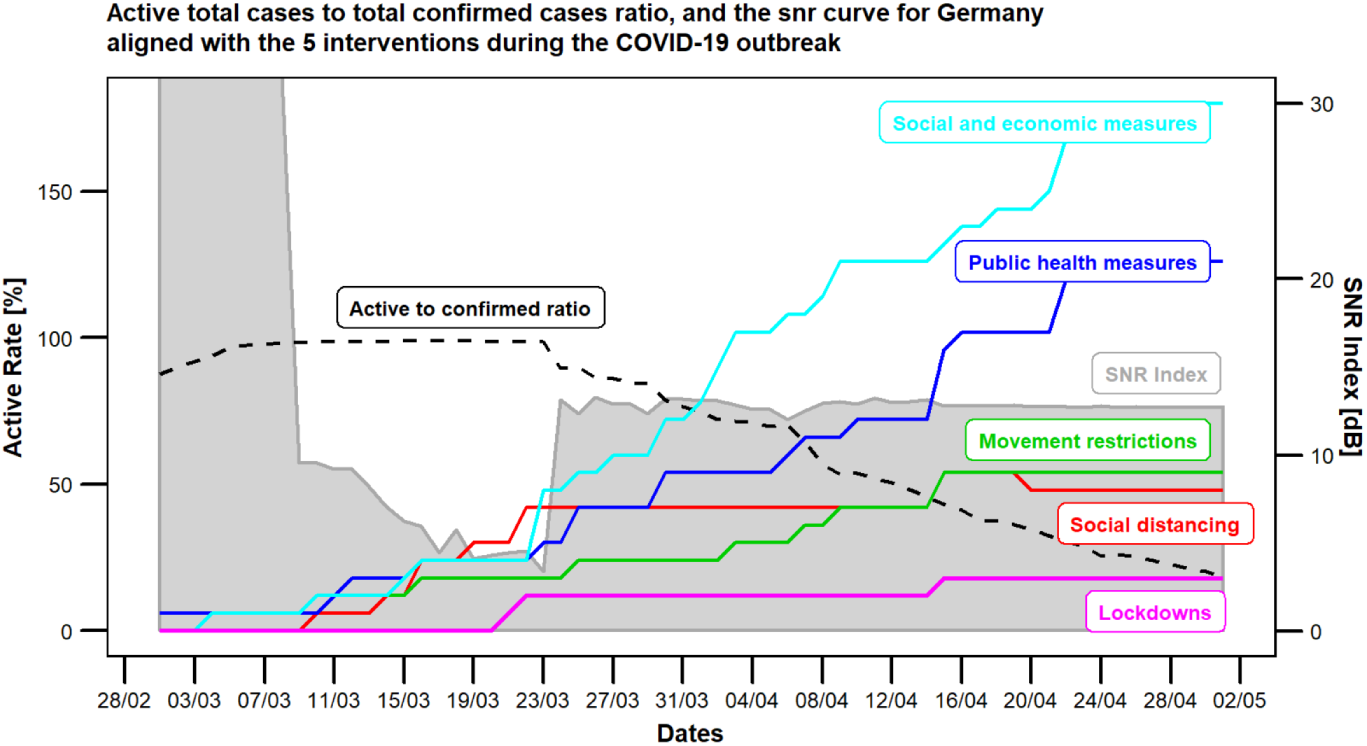
The *snr* Curve and the Active to Confirmed Ratio for Germany along with the Interventions Timeline.

## 5 The impact of testings on death rate, recovered rate and *snr* index

Indubitably, the testing process was one of the critical steps that most countries did not manage to promptly carry out during the early stage of the outbreak due to various reasons. Some of these reasons included but were not limited to: lack of testing kits, not enough health workers, and no adequate facilities to perform the required tests. Nevertheless, some countries thanks to their readiness and strength of their health systems were able to perform the testing with proper pace and volume.

Figures 20 and 21 show clearly, the Germany and the USA were leading the world around the middle of April 2020, in terms of total number of testings. Nevertheless, Germany performed better than the USA on both scales, death rate, *DR* and recover rate, *RR*.

**Figure 20:**
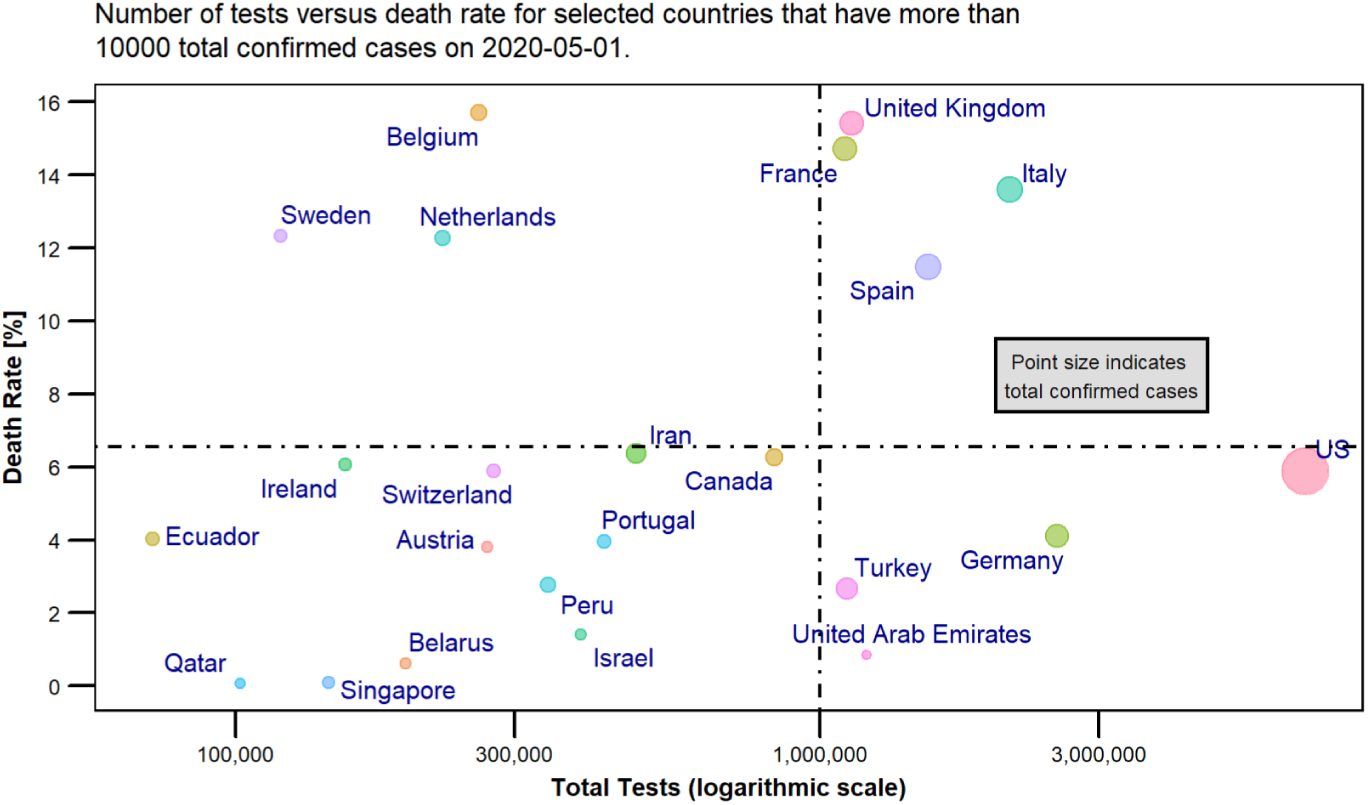
The Relationship between Number of Testings and the Death Rate on 1*^st^* of May 2020.

**Figure 21:**
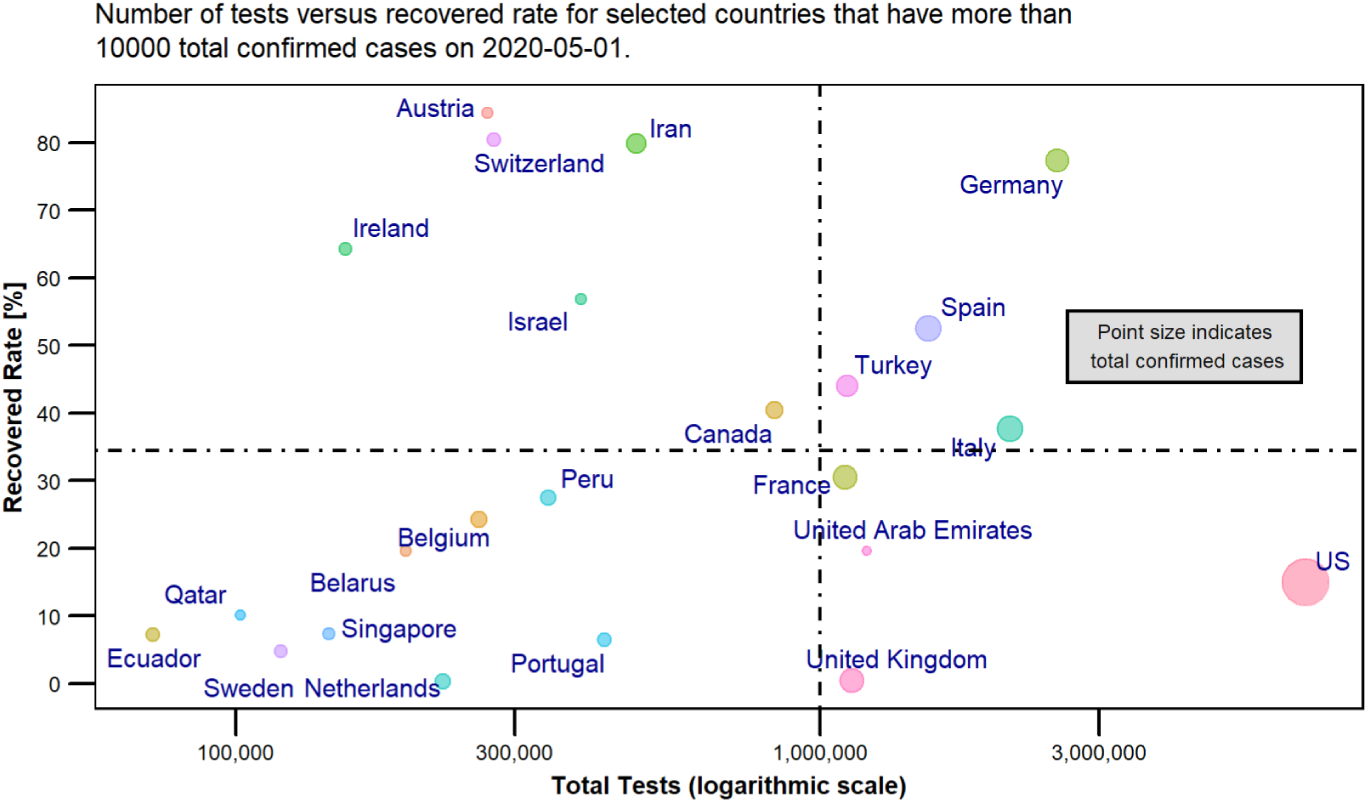
The Relationship between Number of Testings and the Recovered Rate on 1*^st^* of May 2020.

On the other side, it was essential to observe how the newly proposed *snr* index is affected by the total number of tests. Figure 22 shows that only Germany is located in the upper right quadrant indicating its high performance of its health system that managed to carry out a significant number of tests keeping the death rate, *DR* at a lower level than other countries and the recovered rate, *RR* at a higher level. It was of significant importance to validate the application of the proposed *snr* index during the outbreak.

**Figure 22:**
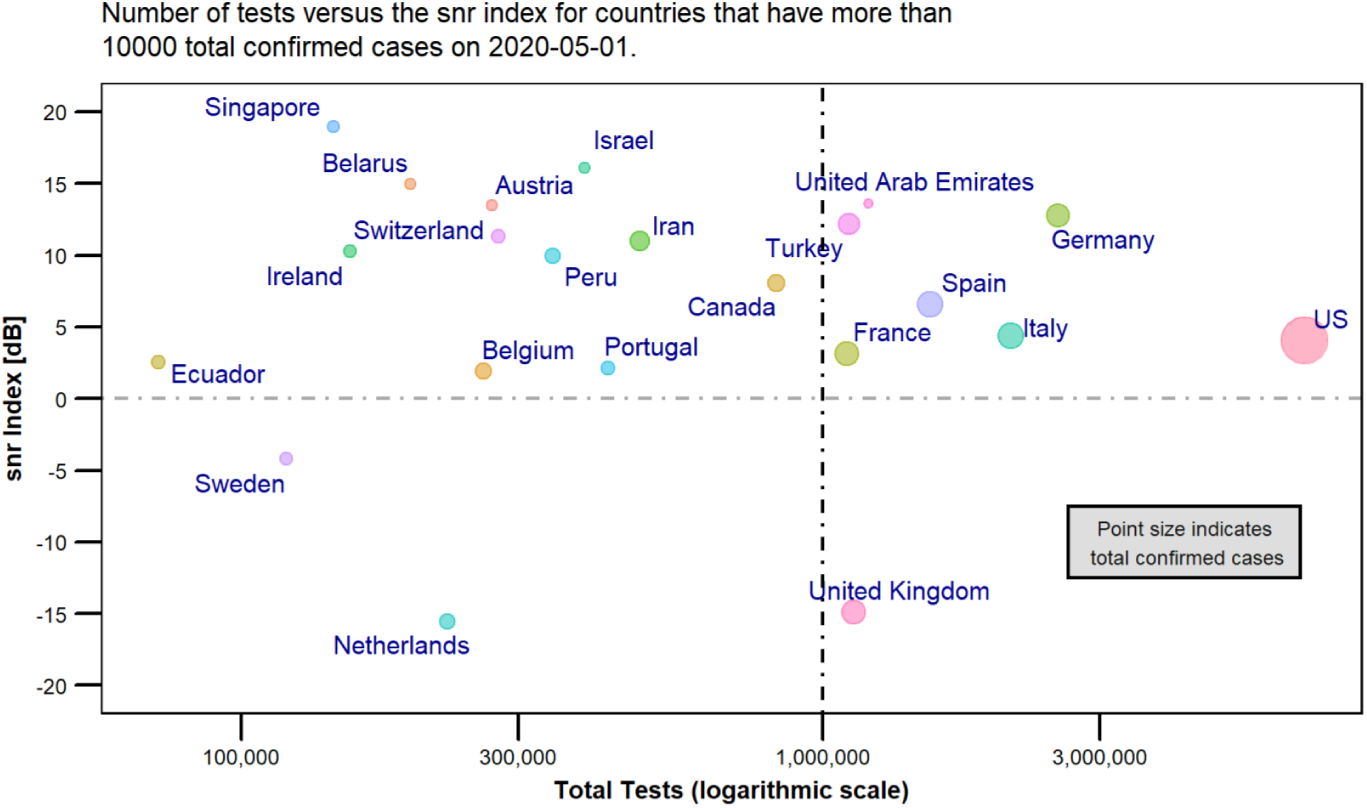
The Relationship between Number of Testings and the Recovered Rate on 1*^st^* of May 2020.

## 6 Conclusion

In this paper the signal to noise ratio, *snr* index was introduced to help assess the effectiveness of the interventions during the COVID-19 pandemic. Moreover, in this paper, it was demonstrated how the *snr* index can provide deeper insights about the efficiency of the health system in terms of capacity and capabilities in dealing with the outbreak of the virus. Although, it was shown that the *snr* index is very useful for decision makers to monitor the effect of their interventions and hence interact promptly with the consequences during crises, nevertheless the index is not useful during pre- and post-crisis. This is due to the nature of the required data which can not be collected except during the outbreak, as per section 3. Nevertheless, this method could be applied not only to a health-related crisis, but also to any type of crises (such as environmental, financial, etc.) in an attempt to monitor promptly the effectiveness of the interventions by the decision makers as well as to assess the efficiency of the operating system. In addition to the proposed *snr* index method, a systems thinking model, with the three dominant feedback loops, for the ongoing COVID-19 crisis was developed to shed light on the critical intervention points that the stakeholders have to consider during any crisis. The model, moreover, works as a template for the future to implore policy makers to act proactively before any such crisis occurs.

## Data Availability

All used data are cited and available under:
https://github.com/CSSEGISandData/COVID-19
and
https://www.worldometers.info/coronavirus/
https://github.com/joachim-gassen/tidy_covid19

https://www.worldometers.info/coronavirus/

https://github.com/joachim-gassen/tidy_covid19

https://ourworldindata.org/grapher/hospital-beds-per-1000-people?tab=chart

## 7 Acknowledgment

A sincere thank you to Dr. Nicola Shaw, Algoma University, Ontario, Canada, for her diligent proofreading of this paper.

## Notes

### Competing Interest Statement

The authors have declared no competing interest.

### Funding Statement

It is not a funded research.

## References

[1] Nicholas LePan. Visual capitalist: Visualizing the history of pandemics, April 2020. URL https://www.visualcapitalist.com/history-of-pandemics-deadliest/.

[2] Max Roser, Hannah Ritchie, Esteban Ortiz-Ospina, and Joe Hasell. Our world in data: Coronavirus pandemic (covid-19), April 2020. URL https://ourworldindata.org/coronavirus#confirmed-covid-19-deaths-by-country.

[3] Oxford University. Coronavirus government response tracker, April 2020. URL https://www.bsg.ox.ac.uk/research/research-projects/coronavirus-government-response-tracker.

[4] ACAPS. Covid19 government measures dataset, April 2020. URL https://www.acaps.org/covid19-government-measures-dataset.

[5] Joachim Gassen. A repository for maintaining covid-19 data in r, April 2020. URL https://github.com/joachim-gassen/tidy_covid19.

[6] Johns Hopkins University Center for Systems Science and Engineering. 2019 novel coronavirus covid-19 (2019-ncov) data repository by johns hopkins csse, April 2020. URL https://github.com/CSSEGISandData/COVID-19.

[7] Dan Chisholm and David B Evans. Improving health system efficiency as a means of moving towards universal coverage. World health report, pages 10-12, 2010.

[8] Pablo Hernandez de Cos and Enrique Moral-Benito. Determinants of health-system efficiency: evidence from oecd countries. International Journal of Health Care Finance and Economics, 14(1):69–93, 2014.

[9] David B Evans, Ajay Tandon, Christopher JL Murray, and Jeremy A Lauer. Comparative efficiency of national health systems: cross national econometric analysis. BMj, 323(7308):307–310, 2001.

[10] Peter Smith. Developing composite indicators for assessing health system efficiency. Measuring Up: Improving the Performance of Health Systems in OECD Countries. Paris: OECD, 2002.

[11] Paul L Delamater, Erica J Street, Timothy F Leslie, Y Tony Yang, and Kathryn H Jacobsen. Complexity of the basic reproduction number (r0). Emerging infectious diseases, 25(1):1, 2019.

[12] Fotios Petropoulos and Spyros Makridakis. Forecasting the novel coronavirus covid-19. PloS one, 15(3):e0231236, 2020.

[13] Worldometers. Covid-19 coronavirus pandemic, April 2020. URL https://www.worldometers.info/coronavirus/.

[14] Elisabeta ROSCA. Stationary and non-stationary time series. The Annals of the” Stefan cel Mare” University of Suceava. Fascicle of The Faculty of Economics and Public Administration, 10:177–186, 01 2011.

[15] Peter M Senge. The fifth discipline: The art and practice of the learning organization. Broadway Business, 2006.

[16] Tom Fiddaman. A community coronavirus model for bozeman, March 2020.

[17] Jeroen Struben. The december 2019 new corona virus (sars-cov-2) outbreak: A behavioral infectious disease policy model. *medRxiv*, 2020.

[18] Hisham Amin and Khaled Wahba. Healthcare performance management model: System dynamics approach. In Proceedings of the 21st International Conference of the System Dynamics Society, pages 20–24, 2003.

[19] Jack B Homer and Gary B Hirsch. System dynamics modeling for public health: background and opportunities. American journal of public health, 96(3):452–458, 2006.

[20] Sawsan Mekki, Manal Abdel Wahed, Khaled K Wahba, and Bassem K Ouda. A system dynamics based model for medical equipment maintenance procedure planning in developing countries. In 2012 Cairo International Biomedical Engineering Conference (CIBEC), pages 104–108. IEEE, 2012.

[21] Patricia Trbovich. Five ways to incorporate systems thinking into healthcare organizations. Biomedical instrumentation & technology, 48(s2):31–36, 2014.

[22] IPAC. Pandemic coronavirus (covid-19), April 2020. URL https://ipac-canada.org/coronavirus-resources.php.

[23] Wikipedia. Signal-to-noise ratio, April 2020. URL https://en.wikipedia.org/wiki/Signal-to-noisejratio.

[24] JoAnn Coleman, Thomas Wrzosek, Robin Roman, John Peterson, and Paul McAllister. Setting system suitability criteria for detectability in high-performance liquid chromatography methods using signal-to-noise ratio statistical tolerance intervals. Journal of Chromatography A, 917(1–2): 23–27, 2001.

[25] Frédéric Lacroix, A Sam Beddar, Mathieu Guillot, Luc Beaulieu, and Luc Gingras. A design methodology using signal-to-noise ratio for plastic scintillation detectors design and performance optimization. Medical physics, 36(11):5214–5220, 2009.

[26] John L Butler and Charles H Sherman. Transducers and arrays for underwater sound. Springer, 2016.

